# O-GlcNAcase dosage variants are associated with neuronal deficits and intellectual disability

**DOI:** 10.1101/2023.11.23.23298195

**Authors:** Florence Authier, Benedetta Attianese, Oscar G. Sevillano-Quispe, Kévin-Sébastien Coquelin, Sergio Galan Bartual, Huijie Yuan, Manuel Peral Vazquez, Iria Esperon-Abril, Andrew T. Ferenbach, Carsten Scavenius, Diane Doummar, Perrine Charles, Cyril Mignot, Boris Keren, Magdalena Krygier, Maria Mazurkiewicz-Bełdzińska, Palle Duun Rohde, Per Qvist, Daan M. F. van Aalten

## Abstract

Rare, high-impact genetic variants affecting genes essential for neurodevelopment represent a major contributing factor to uncharacterised intellectual disabilities. We present the first cases, population-based genetic evidence, an animal model and electrophysiological/functional dissection of pathogenic variants in *OGA*, a key regulator of protein O-GlcNAcylation, associated with cognitive and motor impairments. For a missense variant in the OGA C-terminal HAT domain, a dosage effect in genome-edited mouse embryonic stem cells is associated with impaired neuronal maturation. Mice harbouring this variant display altered brain OGA levels, while maintaining protein O-GlcNAcylation homeostasis. Hippocampal neurons exhibit altered developmental trajectories of neuronal network activity, mechanistically linked to pathways involved in synapse structure and function. We establish *OGA* variants as a cause of intellectual disability and show that partial reduction of OGA levels, even in the absence of global O-GlcNAc perturbation, impairs neuronal maturation and circuit function, highlighting a dosage-sensitive role for OGA in human neurodevelopment.

## Introduction

Protein O-GlcNAcylation is the dynamic posttranslational modification (PTM) in which a single *N*-acetylglucosamine (GlcNAc) moiety is added to serine and threonine residues. In contrast to most other PTMs, O-GlcNAcylation is regulated by only two enzymes: O-GlcNAc transferase (OGT), which catalyses the addition of O-GlcNAc (1), and O-GlcNAcase (OGA), which removes it (2). O-GlcNAcylation regulates a wide range of cellular processes and has been implicated in diverse pathological conditions, including diabetes, neurodegeneration and cancer (3–6). It is essential for embryogenesis and development, as deletion of either *Ogt* or *Oga* (*MGEA5/OGA* in humans) in mice results in lethality and developmental defects (7–11). In the nervous system, O-GlcNAcylation is particularly abundant (2,12,13). It is enriched at synapses (14,15), where numerous proteins important for neurodevelopment and synaptic function have been identified as O-GlcNAc modified - including SynGAP1, Bassoon, Piccolo and NMDA receptors (16,17). Accordingly, pan-neuronal knockout of OGT in mice results in severe neurodevelopmental defects and early neurodegeneration (8), while cell specific deletion of *OGT* in neuronal stem cells disrupts corticogenesis and leads to neuronal stress and apoptosis (18). Similarly, loss of O-GlcNAcylation in forebrain excitatory neurons leads to progressive neurodegeneration and gliosis (19). Together, these findings highlight a critical role for O-GlcNAcylation in brain development, neuronal survival, and function.

Pathogenic variants in *OGT* have been identified in patients with a neurodevelopmental disorder called OGT-linked Congenital Disorder of Glycosylation (OGT-CDG or OGT-ID), characterised by intellectual disability (ID), development delay and dysmorphic features (20–27). While this highlights the importance of O-GlcNAcylation for human neurodevelopment, the precise pathogenic mechanisms remain incompletely understood. Some *OGT* variants directly affect catalytic activity and reduce global O-GlcNAcylation (20–23,28–30), while others located outside the catalytic pocket do not affect global O-GlcNAc levels (20,24,25,27), suggesting the involvement of non-catalytic or indirect mechanisms. Interestingly, cell lines and mice edited to carry OGT-ID variants have shown reduced OGA protein levels, suggestive of a compensatory mechanism to maintain O-GlcNAc homeostasis, yet also itself (reduced OGA) being a possible mechanism contributing to the OGT-ID syndrome (21,22,24,26,28–30).

Common genetic variation in the *OGA* locus have recently been increasingly associated with cognitive traits in large-scale genome-wide association studies (GWAS) (31–36), raising the possibility that primary defects in OGA may themselves be pathogenic. Consistent with a critical role for OGA in the nervous system, loss of OGA levels/activity in animal models is associated with neurodevelopmental consequences. In *Drosophila melanogaster Oga^KO^* flies are viable, yet display locomotion, habituation and synaptic morphology defects (37). In mice, complete loss of OGA activity or expression results in development defects and perinatal lethality (9,11,38), while heterozygous *Oga^+/-^* mice survive but display impaired spatial learning and memory and synaptic plasticity (39). Brain-specific deletion of *Oga* further delays brain development, alters cortical layering and leads to microcephaly accompanied by ventricular enlargement (10). Although these findings establish that reduced OGA dosage can impair brain growth and function in animal models, the contribution of *OGA* pathogenic variants to human neurodevelopmental disorders has not been established.

Here, we strengthen the emerging genetic link between OGA and human cognition by integrating population-based genetic analyses with the identification and functional characterisation of two pathogenic variants in *OGA* in individuals with intellectual disability and developmental delay. Specifically, we identify a patient with a nonsense variant and a patient with a missense variant affecting the C-terminal domain of OGA that is thought to be a “pseudo” histone acetyltransferase (pHAT). Due to the predicted instability of the truncated protein produced in the nonsense variant, our experimental analyses focused on the pHAT-domain missense variant, uncovering effects on OGA protein stability, neuronal maturation, and synaptic function using genome-edited mouse embryonic stem cells, primary neuronal cultures, and a knock-in mouse model. These findings demonstrate that pathogenic OGA variants perturb neuronal and synaptic development, providing the first direct link between *OGA* variants and human neurodevelopmental disease.

## Results

### Genetic variation at the OGA locus influences cognitive performance in the general population

Recent large-scale genome-wide association studies have begun to demonstrate associations between common genetic variation at the *OGA* (also known as *MGEA5*, but consistently referred to as *OGA* here) locus and multiple cognitive traits (31–36). To examine whether genetic variation at this locus influences cognitive performance at the individual level, we leveraged data from the UK Biobank (UKB), which provides rich cognitive phenotyping in a large population-based sample. As UKB does not include a single unified measure of intelligence, we first derived a general cognitive ability (g) score by integrating performance across multiple cognitive tests administered at UKB assessment centres. Using this composite phenotype, we performed a locus-restricted association analysis across a broader genomic interval spanning the *OGA* locus (10q24.32 ± 75 kb). This analysis revealed an association signal across the locus, consistent with patterns reported in large GWAS meta-analyses of cognitive traits, thereby validating the g-score as an appropriate individual-level cognitive phenotype for downstream genetic analyses (**Fig. S1**).

We next examined whether rare protein-altering variants in *OGA* similarly influence cognitive ability. We identified rare coding variants (minor allele frequency < 1 %) mapping to *OGA*, including missense, protein-truncating, splice-region, and in-frame variants, and compared g-scores between carriers and non-carriers using covariate-adjusted models. Carriers of rare coding *OGA* variants exhibited a modest but statistically significant reduction in g-scores relative to non-carriers (**Fig. 1A**, *p* < 0.0142). Given the heterogeneity of variant classes and predicted effects, this pattern suggests that partial perturbation of OGA function or stability can influence cognitive performance at the population level.

**Figure 1:**
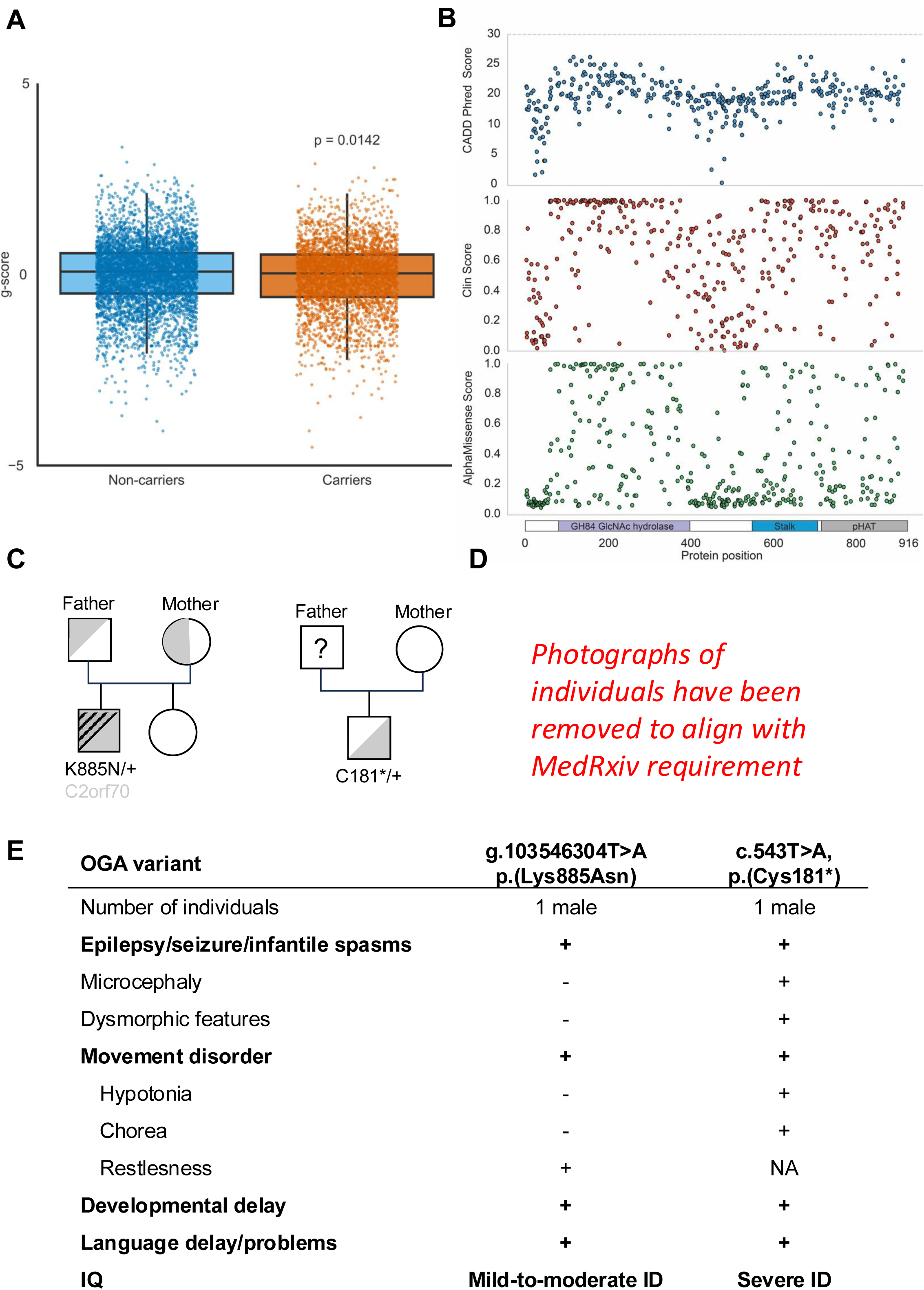
Rare coding OGA variants are associated with general cognitive ability and intellectual disability. **(A)** Boxplot comparing g-score distributions between participants carrying at least one rare (MAF < 1 %) coding variant in the *OGA* gene (n = 3,216) and those participants without rare coding variants in the *OGA* gene (n = 498,075). Individual data dots are overlaid for both groups: a random subsample of 5,000 non-carriers is displayed (blue dots) to avoid excessive overplotting, while all 3,216 individual coding-carriers are shown (orange dots). Statistical significance was assessed using an analysis of covariance (ANCOVA) adjusting for sex, age at recruitment, and UK Biobank assessment centre as covariates (p = 0.0142). **(B)** Multi-panel scatter plot illustrating three independent pathogenicity prediction scores for 419 rare (MAF < 1 %) OGA coding variants plotted against their protein position. The panels show (upper) CADD PHRED score (top panel, blue dots; scale 0–30), (middle) ClinPred score (middle panel, red dots; scale 0–1), and (lower) AlphaMissense pathogenicity score (bottom panel, green dots; scale 0–1). Each dot represents a unique coding variant, plotted at its position in the protein sequence. Pedigrees of the patients with OGA^K885N^ **(C)** and OGA^C181*^ **(D)** variants. **(E)** Table recapitulating the clinical findings in patients affected with the OGA^K885N^ and OGA^C181*^ variants. **(F)** Clinical photographs of patient with OGA^C181*^ variant showing dysmorphic features.

To place these findings in a structural context, we examined the distribution of rare coding variants across the OGA protein in relation to predicted pathogenicity scores (**Fig. 1B**). Variants with higher predicted pathogenicity were more frequently observed within the N-terminal GlcNAc hydrolase catalytic domain, which contains the core enzymatic machinery, as well as within a region of the central stalk domain that is part of the OGA dimerization interface. In contrast, fewer high-scoring variants were observed in the distal pHAT domain, and these did not form a distinct cluster. Together, these observations indicate that predicted damaging population variation is unevenly distributed across OGA, preferentially affecting structurally and functionally constrained regions of the protein.

### Identification of OGA variants in two unrelated individuals with intellectual disability, motor impairment and epilepsy

To bridge population-level genetic associations with clinical neurodevelopmental phenotypes, we investigated whether rare coding variants in *OGA* occur in individuals with ID. We identified two cases with ID carrying variants in *OGA*.

Proband I, referred in late adolescence for etiological investigation of ID, was born to healthy unrelated parents following an unremarkable pregnancy and perinatal course. Birth occurred at 36 weeks of amenorrhea with normal parameters: weight 3440 g, length 49 cm, OFC 36 cm, and Apgar scores of 10 at both 1 and 5 minutes. There were no neurodevelopmental concerns during the first year of life. The patient was in the 25^th^ percentile age range for sitting alone, 10^th^ percentile for walking independently (percentile range ages are defined according to World Health Organization (40)), and spoke disyllabic words from the age of 12 months. Infantile spasms, a rare type of epilepsy, developed after the first year of life. Early motor and language development was mildly delayed, and infantile spasms developed after the first year of life, with EEG recordings showing hypsarrhythmia, while brain MRI was normal. Seizures responded well to treatment, and no further seizure types were observed until early adulthood. The parents noted psychomotor retardation and restlessness after the onset of epilepsy. However, language, motor and social skills progressed from the start of his middle childhood (defined as 6-12 years). Cognitive assessment during this period indicated mild to moderate ID (WISC-IV, FSIQ of 54, VCI 57, PRI 69, WMI 67, PSI 71), with a heterogeneous cognitive profile (WAIS, VCI 59, PRI 84, WMI 66, SPI 78) persisting into adulthood. Finally, one seizure occurred at the beginning of his twenties, with no further EEG abnormalities. Whole exome sequencing identified a heterozygous missense mutation in the pHAT domain of *OGA* (GRCh37: Chr10:g.103546304T>A p.(Lys885Asn). An additional homozygous variant of unknown significance in a splicing region of the *C2orf70* gene (NM_001105519.2:c.462-7C>G) was identified, but segregation analysis demonstrated parental inheritance (**Fig. 1C**). Population constraint metrics indicate that *OGA* is highly intolerant to both missense and loss-of-function variation, whereas *C2orf70* appears tolerant to genetic variation (41). Consistent with this, C2orf70/FAM166C protein was not detected in mouse embryonic stem cells or embryonic brain tissue, suggesting limited relevance for prenatal neurodevelopment (**Fig. S2**). Together, these findings support *OGA* as the primary candidate gene in this individual.

Proband II, evaluated in middle childhood (defined as 6-12 years) presented with severe global developmental delay, epilepsy, hypotonia (**Fig. 1D**), generalized chorea (**Video S1**), and dysmorphic features. He is the single child of healthy unrelated individuals with an uneventful family history. Birth occurred through normal vaginal delivery at 39 weeks of uncomplicated gestation with birth weight of 3600 g and Apgar score of 5. After birth, problems with breathing occurred and the patient spent two weeks in intensive care but no formal documents are available from that time. Developmental milestones were markedly delayed in all areas. He started to sit independently from the age of 1,5 years oand stand from the age of 4 years. From the newborn period the patient showed hypotonia and he presented involuntary movements from the first year of life. From 3-4 year of age, the patient presented with restricted language skills using simple words (yes, no, mama) and not forming sentences. The mother noted the presence of epileptic seizures from the first months of life in the form of upper eye deviations and from about three months of age he has been receiving valproic acid. He is formally diagnosed with severe intellectual disability. From the age of 6 years, he received a low dose of valproate (2× 100 mg per day). Seizures in the form of “sudden head flexion” (head drop) were observed every few days. EEG recording showed focal epileptic discharges. Neurological examination at that time was notable for microcephaly (HD 50 cm, below 3rd percentile at six years old), dysmorphic features including high-arched palate, low-set ears, micro-retrognathia, pectus excavatum and pes plano-valgus and generalized muscular hypotonia (**Fig. 1D**). The patient also presented generalized chorea allowing walking only with assistance (**Video S1**). Brain MRI showed no structural brain abnormalities and ultrasound examination of the abdomen was normal.

Ophthalmological evaluation showed post-traumatic corneal scars, otherwise normal. The dose of valproate was increased and was later associated with clobazamum with a positive outcome. The patient is currently seizure-free for several months. Recent evaluation in his middle childhood (defined as 6-12 years) was notable for prominent hyperkinetic movement disorder. There were no ataxia or pyramidal signs, tendon reflexes were present, normal and symmetric. Although the patient was able to independently walk several steps, he was wheelchair dependent. The patient was only able to speak single words. Routine laboratory investigations were normal. Whole exome sequencing identified a heterozygous nonsense variant in *MGEA5/OGA* (GRCh37: Chr:10:103567596, NM_012215.5, c.543T>A, p.(Cys181*)). The variant was absent in the mother, and paternal DNA was unavailable (**Fig. 1C**). The predicted truncation removes most of the functional part of the OGA protein, with this patient effectively being heterozygous for functional OGA.

A summary of clinical features for both individuals is provided in **Fig 1E**. Taken together, these findings identify two unrelated individuals with ID carrying variants in *OGA*, including one truncating and one missense variant affecting the pHAT conserved domain. These variants are henceforth referred to as OGA-ID variants.

### OGA-ID variants imply loss of function

Structural analyses suggest potential mechanisms by which the two OGA-ID variants may lead to impaired OGA function and/or stability. Inspection of the published crystal and cryoEM structures of OGA (42–45) suggests that the Cys181* nonsense variant leads to truncation of the glycoside hydrolase catalytic core and complete loss of the pHAT domain (**Fig. 2A,B**). The topology of the OGA catalytic core adopts a (β/α)_8_-barrel fold, with the key catalytic residues Asp 174 and Asp 175 located on a loop after the β4 repeat (**Fig. 2B**). The putative expressed truncated protein has a β4/α3 topology, which is likely insufficient to form a stable, soluble protein. Furthermore, OGA binds and orients the GlcNAc moiety from the O-GlcNAcylated proteins using residues distributed along the full TIM barrel (Lys98, Gly67, Asp285 and Asn280) (**Fig. 2B**). Thus, OGA Cys181*is unlikely to be able to bind O-GlcNAc or to retain any catalytic activity.

**Figure 2:**
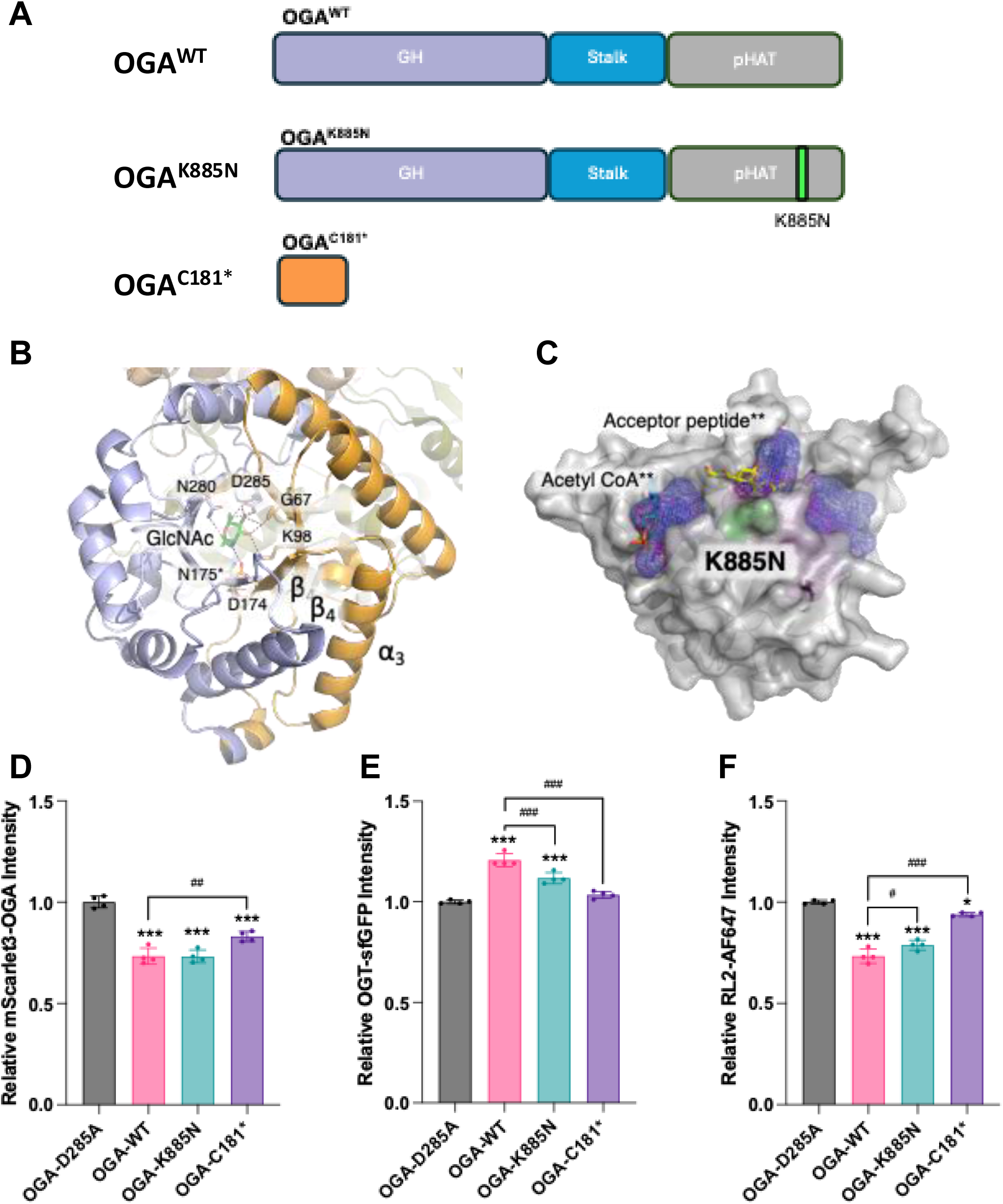
Structural and functional consequences of the OGA-ID variants. **(A)** Schematic organisation of the modular OGA protein and the OGA^C181*^ and OGA^K885N^ variants. For clarity, each module/mutation is coloured differently. **(B)** Detailed view of the OGA TIM barrel (PDB: 5vvx). The predicted expressed polypeptide (1–180) is coloured in orange while the non-expressed OGA (181 to 715) is shown as a light blue cartoon. GlcNAc moiety is coloured in green, and the interacting residues are labelled and shown as sticks. (*) Note that in the PDB structure, the Asp 175 was mutated to Asn in order to generate an inactive OGA and promote the complex with GlcNAc (92). **(C)** Surface representation of the pHAT^K885N^ using as template the PDB (93). (**) Similarly, the acetyl-CoA and the acceptor peptide were obtained from the tGCN5 structure while the cavities were detected with the PyMOL plugin CavitOmix. The mutation site is coloured in green. **(D)**, OGT-sfGFP **(E)** and O-GlcNAc **(F)** in dual-tagged mESCs measured by FACS. The catalytically inactive OGA^D285A^ mutant served as a control and baseline for normalising all signals. Each dot represents an independent replicate; experiments were performed on different days using cells of varying passage numbers. Data are represented as median ± SEM (n = 4-6). Significance using ordinary one-way ANOVA shown as ns (not significant, *p* > 0.05), * (*p* < 0.05), ** (*p* < 0.01), *** (*p* < 0.001).

The K885N OGA-ID variant results in an amino acid substitution in the C-terminal pHAT protein domain (**Fig. 2A, C**). This domain possesses a fold related to the Gcn5 family of acetyltransferases (46), but with two additional helices, *α*_3_ and *α*_4_ inserted between strands *β*_3_ and *β*_4_, reducing the size of the Gcn5 family conserved peptide binding cleft (**Fig. 2C**) (42,43). Comparison of the recently published human OGA pHAT model (42) with the structure of a Gcn5 protein from *Tetrahymena thermophila* (tGcn5) in complex with an acceptor peptide (RMSD = 3.9 Å over 80 Ca atoms) (46) places the peptide inside this cleft (**Fig. 2C**). Interestingly, the OGA K885N mutation is placed at the C-terminus of the a7 helix at one of the cleft entrances and in *t*Gcn5, the residue in the equivalent position (N163) was suggested to participate in substrate stabilisation (46). Experiments to measure effects of the K885N mutation on OGA stability (by differential scanning fluorimetry (DSF, **Fig. S3A**) and on the catalytic O-GlcNAcase activity (steady state kinetics with a pseudo substrate, **Fig. S3B**) did not reveal significant differences between wild type and mutant forms of the OGA protein. Taken together, structural analyses are consistent with the OGA-ID variants leading to either complete loss of the pHAT domain or to altered pHAT function.

### OGA-ID variants modulate O-GlcNAc homeostasis

To assess the effects of the OGA-ID variants on OGA function and O-GlcNAc homeostasis, we exploited a recently reported triple fluorescence stem cell line (3FmESCs) that allows simultaneous quantification of OGT, OGA and O-GlcNAc levels (47) after overexpression of OGA/OGT variants, exploiting the previously reported feedback mechanisms that maintain O-GlcNAc homeostasis (**Fig. S4A**). As a negative control, we first investigated the ten most common OGA variants in healthy individuals sourced from gnomAD v.4.0.0 (41). When introducing these variants into the 3FmESCs, the cells exhibited a compensatory reduction in OGA-mScarlet3 fluorescence (**Fig. S4B**) and an increase of OGT-sfGFP fluorescence (**Fig. S4C**) compared to the OGA^D175N^ catalytically inactive mutant that did not induce such changes. The changes were comparable to those observed with wild type OGA (OGA^WT^) transfection, suggesting unaltered OGA function of the non-pathogenic gnomAD OGA variants. We next assessed the effects of the OGA^K885N^ and OGA^C181*^ OGA-ID variants on O-GlcNAc homeostasis. For the OGA^C181*^ variant a significantly different response compared to the OGA^WT^ protein was observed (**Fig. 2D**). Introducing the OGA^K885N^ variant also led to a significant increase of OGT-sfGFP fluorescence and a significant reduction in RL2 signal, albeit to a lesser extent compared to OGA^WT^ (**Fig. 2E, F**), consistent with altered modulation of O-GlcNAc homeostasis. In contrast, no change in either OGT-sfGFP fluorescence or RL2 signal was observed with the OGA^C181*^ variant (**Fig. 2E, F**), consistent with a complete loss of OGA function. Taken together, these data indicate that OGA-ID variants differentially affect O-GlcNAc homeostasis relative to wild type OGA.

### Increased OGA turnover and neuronal maturation deficit in Oga^K885N^ mESCs

We next generated mESCs carrying the OGA^K885N^ mutation using CRISPR/Cas9 genome editing. mESC lines of the three genotypes *Oga*^+/+^, *Oga*^K885N/+^ and *Oga*^K885/K885N^ were validated by whole genome sequencing to exclude off-target effects and used for further analysis. We first investigated the effect of the K885N variant on O-GlcNAc homeostasis. *Oga*^+/+^ and *Oga*^K885N/+^ mESCs showed similar OGA protein levels while *Oga*^K885N/K885N^ cells showed a significant reduction (**Fig. 3A, B**). This reduction was not associated with changes in OGT protein levels (**Fig. 3A, C**) or global O-GlcNAc levels (**Fig. 3A, D**) in any of the three genotypes. We also observed similar *Oga* and *Ogt* mRNA levels across the three genotypes (**Fig. 3E, F**).

**Figure 3:**
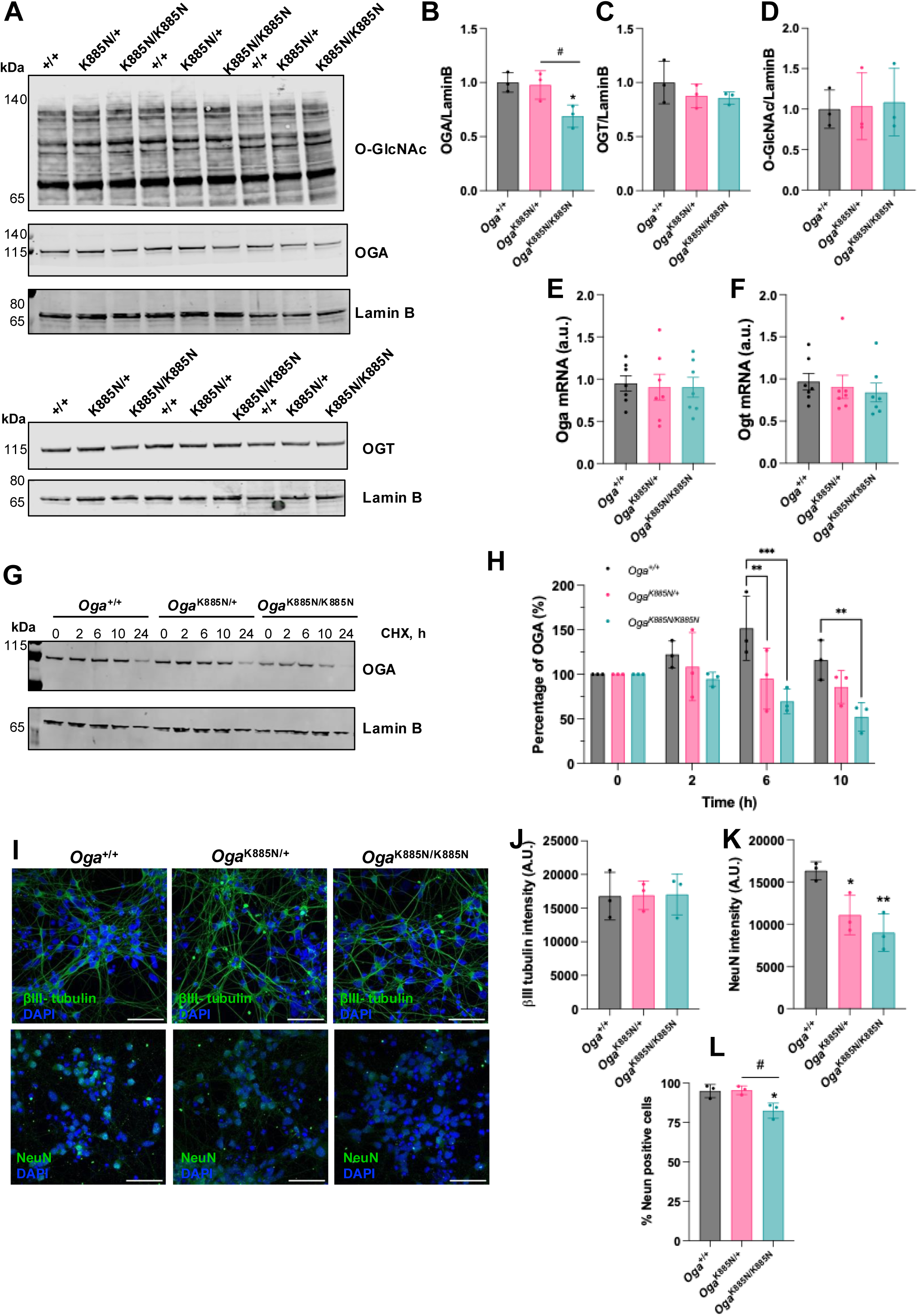
The OGA-K885N mutation causes reduction of OGA protein levels due to increased turnover. Data are represented as mean ± SD, of minimum n = 3 independent experiments for all genotypes. Significance using One-way Anova with Tukey multiple comparison is shown as * *p* < 0.05 and ** *p* < 0.01**. (A)** Western blot of O-GlcNAc and OGA protein levels in *Oga*^+/+^, *Oga*^K885N/+^ and *Oga*^K885N/K885N^ mESCs. Lamin B antibody was used as loading control. Quantification of OGA **(B)**, OGT **(C)** and O-GlcNAcylated **(D)** protein levels from the Western blot in panel A. Quantification of the *Oga* **(E)** and *Ogt* **(F)** mRNA levels in *Oga*^+/+^, *Oga*^K885N/+^ and *Oga*^K885N/K885N^ mESCs. **(G)** Western blot of OGA protein levels during CHX assay in *Oga*^+/+^, *Oga*^K885N/+^ and *Oga*^K885N/K885N^ mESCs for the indicated time points. Lamin B antibody was used as loading control. **(H)** Percentage of OGA protein levels quantified from panel G. **(I)** Representative fluorescent microscopy images of βIII tubulin and NeuN expression in *Oga*^+/+^, *Oga*^K885N/+^ and *Oga*^K885N/K885N^ mutant cells after 18 days of neurodifferentiation. Magnification 40x, 50 μm scale bar. Quantification of mean intensity of βIII tubulin **(J)**, NeuN **(K)** signals and number NeuN positive cells (**L**) from panel I.

To investigate whether the reduction in OGA protein levels is caused by increased OGA turnover, *Oga*^+/+^, *Oga*^K885N/+^ and *Oga*^K885N/K885N^ mESCs were treated with the protein synthesis inhibitor cycloheximide (CHX) and OGA protein levels were assessed over a 10 h period. Both *Oga*^K885N/+^ and *Oga*^K885N/K885N^ cells showed an early decrease of OGA protein levels at 6 h, progressing to 26 % and 55 % reduction, respectively, after 10 h (**Fig. 3 G, H**).

We next evaluated the pluripotency state of the mESCs by both qPCR and immunostaining. *Oga*^+/+^, *Oga*^K885N/+^ and *Oga*^K885N/K885N^ mESCs showed no significant difference in *nanog* or *sox-2* protein (**Fig. S5A-C**) and mRNA levels (**Fig. S5 D, E**), suggesting that the K885N mutation does not affect pluripotency or self-renewal. Upon 18 days of neuronal differentiation, all three mESC lines formed neuron-like structures with neurites and cell bodies, and similar levels of βIII-tubulin were detected suggesting comparable neuronal differentiation between the three genotypes (**Fig. 4I, J**). However, both *Oga*^K885N/+^ and *Oga*^K885N/K885N^ mESC-derived neurons showed decreased levels of the post-mitotic marker NeuN, suggesting a neuronal maturation defect (**Fig. 4I, K**). In Oga^K885N/K885N^ mESC-derived neurons, the reduction of NeuN expression was associated with a lower number of NeuN positive cells compared to both *Oga*^+/+^ and *Oga*^K885N/+^ genotypes (**Fig. 4L**). Taken together, these data show that the *Oga*^K885N^ mutation primarily reduces OGA levels through increased protein turnover and that, while neuronal differentiation can be induced in *Oga*^K885N^ mutant mESCs, neuronal maturation is impaired.

**Figure 4:**
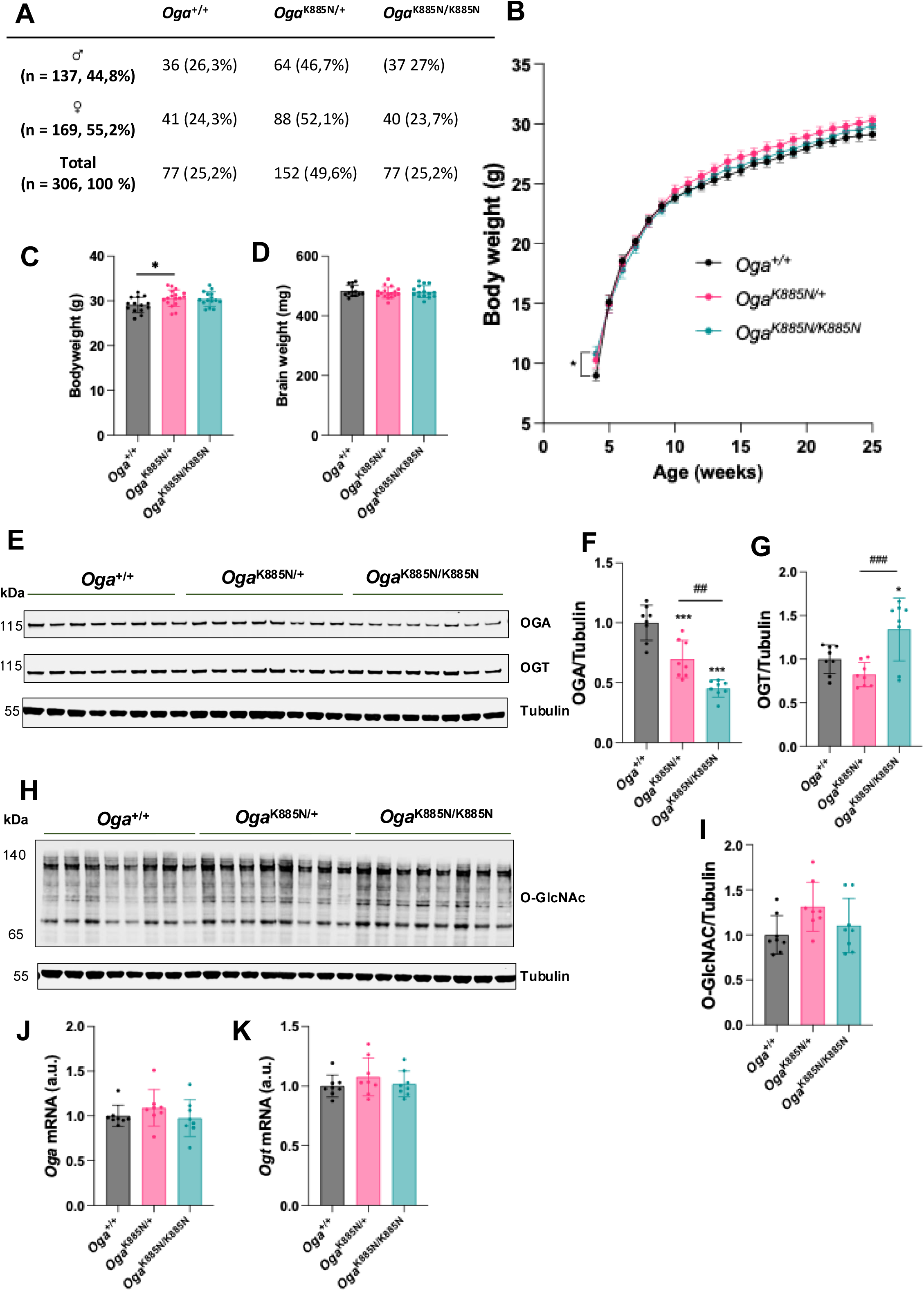
*Oga*^K885N^ mice show decreased levels of OGA protein levels in brain. Data are represented as mean ± SEM. Significance using a *t*-test or One-way Anova with Tukey multiple comparison is shown as * *p* < 0.05 and ** *p* < 0.01. **(A)** Table representing mendelian ratio of *Oga*^K885N^ animals generated from breeding pairs of female *Oga*^K885N/+^ and male *Oga*^K885N/+.^ **(B)** Body weight of male *Oga*^+/+^(n = 14), *Oga*^K885N/+^ (n = 17) and *Oga*^K885N/K885N^ (n = 15) mice from 4 weeks to 25 weeks old. Body weight **(C)** and brain weight **(D)** of 25 weeks old male *Oga*^+/+^ (n = 14), *Oga*^K885N/+^ (n = 17) and *Oga*^K885N/K885N^ (n = 15) mice. **(E)** Western blot of OGA and OGT protein levels in hippocampus tissues from 25 weeks old male *Oga*^+/+^, *Oga*^K885N/+^ and *Oga*^K885N/K885N^ mice (n = 8 per genotype). Tubulin antibody was used as loading control. Quantification of OGA **(F)** and OGT **(G)** protein levels from the Western blot in panel E. **(H)** Western blot of O-GlcNAc proteins in hippocampus tissues from 25 weeks old male *Oga*^+/+^, *Oga*^K885N/+^ and *Oga*^K885N/K885N^ mice (n = 8 per genotype). Tubulin antibody was used as loading control. **(I)** Quantification of O-GlcNAcylated protein levels from the Western blot in panel H. Quantification of the *Oga* **(J)** and *Ogt* **(K)** mRNA levels in hippocampus tissues from 25 weeks old male *Oga*^+/+^, *Oga*^K885N/+^ and *Oga*^K885N/K885N^ mice (n = 8 per genotype).

### Reduction of OGA protein levels in Oga^K885N^ mice

To dissect the effects of the OGA^K885N^ variant *in vivo*, we introduced this variant in mice using a previously described CRISPR/Cas9 editing approach (28). After zygotic injection of the editing reagents, DNA from offspring mice was genotyped and sequenced to confirm the presence of the K885N mutation. Mendelian analysis revealed normal gender and genotype distributions (**Fig. 4A**). Only male animals were used for all subsequent experiments. Both *Oga*^K885N/+^ and *Oga*^K885N/K885N^ mice develop similarly to *Oga*^+/+^ mice as assessed by body and brain weight (**Fig. 4B-D**).

We then measured O-GlcNAc homeostasis in the hippocampus from *Oga*^K885N^ mice by western blot and qPCR. OGA protein levels were reduced in *Oga*^K885N/+^ and *Oga*^K885N/K885N^ mice compared to *Oga*^+/+^ mice (**Fig. 4E, F**). While OGT protein levels were increased in *Oga*^K885N/K885N^ mice compared to *Oga*^K885N/+^ and *Oga*^+/+^ mice (**Fig. 4E, G**), global O-GlcNAc levels were unchanged in all three genotypes (**Fig. 4H, I**). The changes in OGA/OGT protein levels were not associated with differences in *Oga* and *Ogt* transcript levels between genotypes (**Fig. 4J, K**), suggesting post-translational mechanisms. Taken together these data are consistent with our observation in mESCs and indicate that the hippocampus maintains global O-GlcNAc levels despite reduced OGA protein levels.

### Oga^K885N^ mice show mild behavioural deficits

To investigate whether the K885N variant alters behaviour *in vivo*, we assessed locomotion, anxiety-related behaviour, repetitive behaviour, and learning and memory across *Oga*^+/+^, *Oga*^K885N/+^ and *Oga*^K885N/K885N^ mice. General locomotor activity and motor coordination were first assessed to exclude gross neurological impairments. No difference in total distance travelled in the open field was observed in any of the three genotypes (**Fig. 5A, B**). We then assessed balance and coordination using the static rod tests. Both *Oga*^K885N/+^ and *Oga*^K885N/K885N^ mice completed the required t-turns at a speed comparable to *Oga*^+/+^ control mice (**Fig. 5C**), suggesting conserved balance and coordination in *Oga*^K885N^ animals.

**Figure 5:**
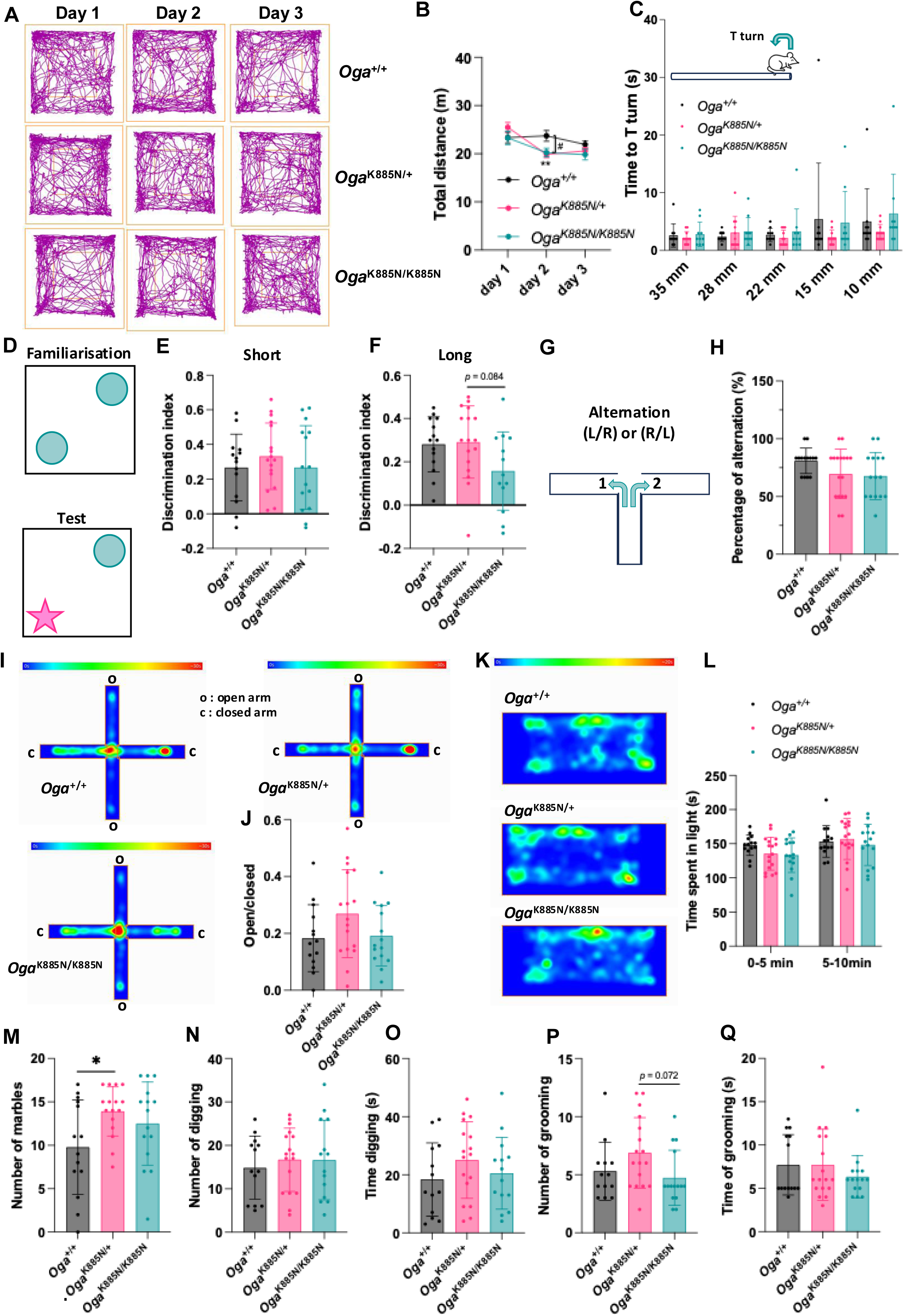
*Oga*^K885N^ mice show mild compulsive behaviour. For all behaviour paradigms, Two-way ANOVA (Alpha, 0.05) was used, followed by Tukey’s column comparisons. Significance is shown as * *p* < 0.05 and ** *p* < 0.01. Data are represented as mean ± SD**. (A)** Representative tracking plot over three consecutive days of a male *Oga*^+/+^, *Oga*^K885N/+^ and *Oga*^K885N/K885N^ mice in the open field arena. **(B)** Distance travelled over three consecutive days of male *Oga*^+/+^ (n = 14), *Oga*^K885N/+^ (n = 17) and *Oga*^K885N/K885N^ (n = 15) mice in the open field arena. **(C)** Time to t-turn of male *Oga*^+/+^ (n = 14), *Oga*^K885N/+^ (n = 17) and *Oga*^K885N/K885N^ (n = 15) mice during the static rods test. **(D)** Representative image of the Novel Object Recognition (NOR) test. During the familiarization phase, mice are free to explore an arena with two identical objects. During the test phase, mice were free to explore the same arena where one of the familiar objects has been replaced by a novel object to assess short (90 min) and long (24 h) memory. Discrimination index of male *Oga*^+/+^ (n = 14), *Oga*^K885N/+^ (n = 17) and *Oga*^K885N/K885N^ (n = 15) mice during the short term **(E)** and long term **(F)** NOR tests. **(G)** Schematic of the T-maze paradigm. Mice are free to explore both arms for 7 additional trials. Each choice was recorded. **(H)** Percentage of correct alternation (L-R/R-L sequences) of male *Oga*^+/+^ (n = 14), *Oga*^K885N/+^ (n = 17) and *Oga*^K885N/K885N^ (n = 15) mice during the T-maze test. **(I)** Representative heat maps of male *Oga*^+/+^, *Oga*^K885N/+^ and *Oga*^K885N/K885N^ mice in the elevated plus maze (EPM). **(J)** Ratio between time spent in open and close arms by male *Oga*^+/+^ (n = 14), *Oga*^K885N/+^ (n = 17) and *Oga*^K885N/K885N^ (n = 15) mice in the EPM. **(K)** Representative heat maps of male *Oga*^+/+^, *Oga*^K885N/+^ and *Oga*^K885N/K885N^ mice in the light compartment during the dark-light paradigm (D/L). **(L)** Time spent of male *Oga*^+/+^ (n = 14), *Oga*^K885N/+^ (n = 17) and *Oga*^K885N/K885N^ (n = 15) mice in the light compartment during the dark-light paradigm (D/L). **(M)** Number of buried marbles by male *Oga*^+/+^ (n = 14), *Oga*^K885N/+^ (n = 17) and *Oga*^K885N/K885N^ (n = 15) mice during the marble test. Number of digging events **(N)** and time spent digging **(O)** by male *Oga*^+/+^ (n = 14), *Oga*^K885N/+^ (n= 17) and *Oga*^K885N/K885N^ (n = 15) mice during 3 min observation. Number of self-grooming events **(P)** and time spent self-grooming **(Q)** by male *Oga*^+/+^ (n = 14), *Oga*^K885N/+^ (n = 17) and *Oga*^K885N/K885N^ (n = 15) mice during 3 min observation.

We next investigated the effect of the *Oga*^K885N^ variant on memory. We assessed novelty-associated memory using the Novel Object Recognition (NOR) paradigm (**Fig. 5D**). While after both 90 min (**Fig. 5E**) and 24 h (**Fig. 5F**) intervals, *Oga*^K885N/+^ and *Oga*^K885N/K885N^ mice showed different discrimination indices compared the *Oga*^+/+^ mice, these differences are not significant given the experimental variation. We did not observe significant differences in percentage of spontaneous alternation during in T-maze tests for all three genotypes (**Fig. 5G, H**), suggesting proper working spatial memory.

Anxiety-like behaviours were then assessed during Elevated Plus Maze (EPM) and dark-light paradigm tests. No difference in open to closed time ratios were observed between all three genotypes during the EPM test (**Fig. 5I, J**). Similarly, *Oga*^K885N/+^ and *Oga*^K885N/K885N^ mice spent comparable time in the light compartment compared to *Oga*^+/+^ mice during the dark-light paradigm test (**Fig. 5K, L**). These findings suggest the absence of light/height-induced anxiety-like behaviours in the *Oga*^K885N^ mice.

Finally, we assessed compulsive and repetitive behaviour. Both *Oga*^K885N/+^ and *Oga*^K885N/K885N^ mice showed an increased number of buried marbles in a marble burying test compared to *Oga*^+/+^ mice (**Fig. 5M**). Although no difference in digging events (**Fig. 5N**) was observed between all three genotypes, *Oga*^K885N/+^ mice spent more time digging compared to both *Oga*^+/+^ and *Oga*^K885N/K885N^ animals (**Fig. 5O**), although not significantly. The number of self-grooming events and time spent self-grooming were similar between the three genotypes (**Fig. 5P, Q**). Taken together, these findings suggest mild levels of compulsive behaviour in *Oga*^K885N^ mice.

### Hippocampal proteomic changes suggest perturbed synaptic molecular pathways

To get insight into the potential processes affected by the K885N variant, we performed proteomics analysis on hippocampal tissues of *Oga*^+/+^, *Oga*^K885N/+^ and *Oga*^K885N/K885N^ mice using label-free quantification. A total of 4713 proteins were identified with a 1 % false discovery rate. Principal Component Analysis (PCA) showed that *Oga*^+/+^, *Oga*^K885N/+^ and *Oga*^K885N/K885N^ mice clustered in three separate groups suggesting consistently different proteome profiles between all three genotypes (**Fig. 6A**). We then investigated the proteins that were deregulated in *Oga*^K885N/+^ and *Oga*^K885N/K885N^ compared to wild type mice based on defined cut-off criteria (*p ≤ 0.05)* **(Fig. 6B, C**). Among the deregulated proteins, we observed consistent downregulation of OGA in both *Oga*^K885N/+^ and *Oga*^K885N/K885N^ genotypes compared to wild type (**Fig. 6B, C**), aligning with our western blot analysis (**Fig. 4E, F)**. We then focused on identifying proteins that changed similarly in *Oga*^K885N/+^ and *Oga*^K885N/K885N^ mice when compared to wild type control. We identified a subset of 285 proteins that were either upregulated or downregulated in both *Oga*^K885N/+^ and *Oga*^K885N/K885N^ mice (**Fig. 6D**). Cellular enrichment analysis revealed that these proteins mainly localised in myelin, mitochondrial and synaptic compartments (**Fig. 6E**). Gene Ontology (GO) analysis of these proteins highlighted involvement in key biological processes such as mTORC1 signaling (GO:1903432), nucleoside metabolism (GO:0006753), protein translation (GO:0006412) and folding (GO:0006457) (**Fig. 6E**). In accordance with the abundance of deregulated synaptic proteins, pathways regulating synapse organisation (GO:0050807) and neuron projection development (GO:0010975) were also significantly enriched (**Fig. 6E**). Taken together these data show that the K885N mutation is associated with perturbed molecular pathways suggesting possible synaptic dysfunction.

**Figure 6:**
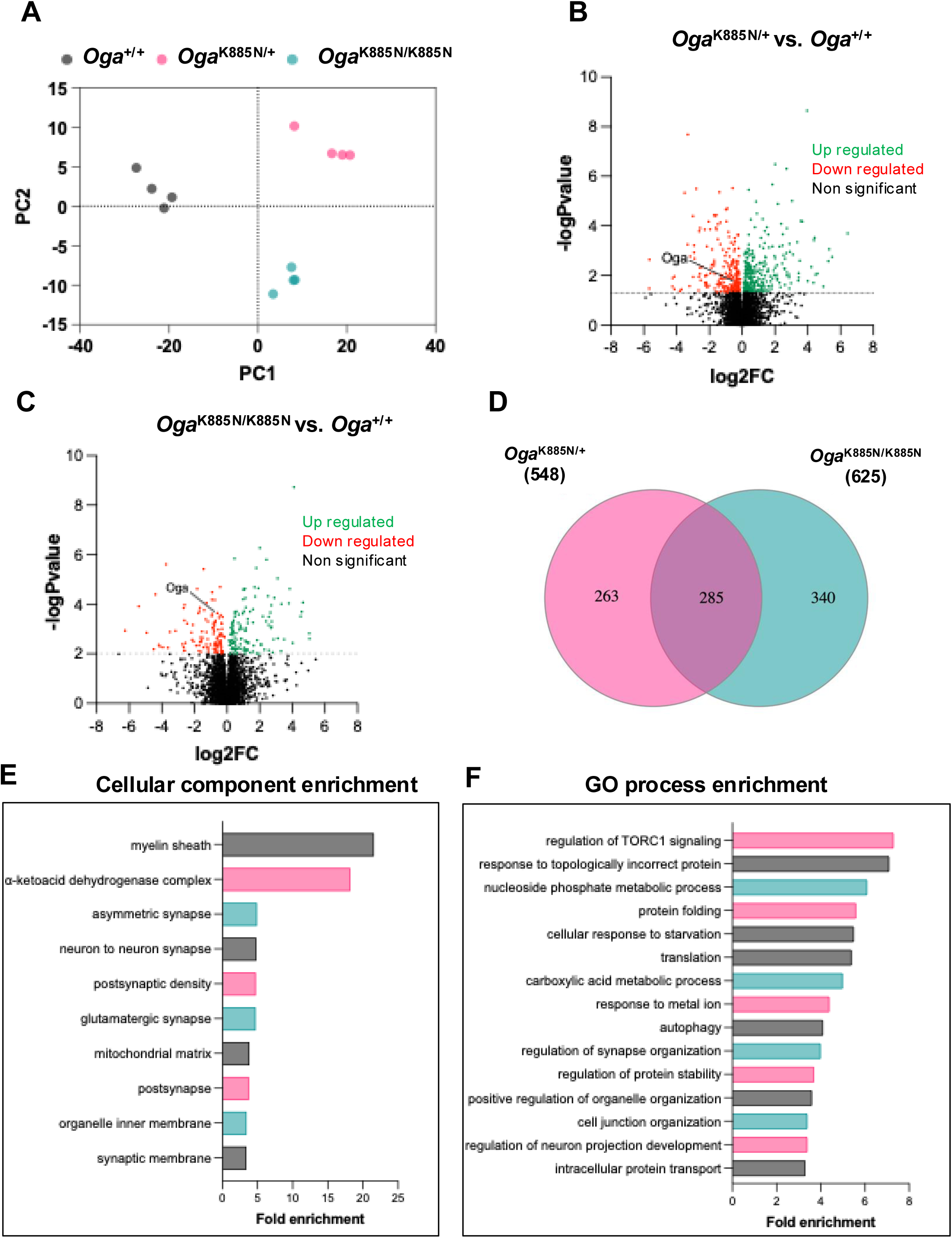
Quantitative proteomics analyses revealed distinctly perturbed synaptic molecular pathways in hippocampus of the *Oga*^K885N^ mice. **(A)** PCA biplot of PC1 and PC2 of the normalised protein abundances in *Oga*^+/+^, *Oga*^K885N/+^ and *Oga*^K885N/K885N^ mice. Volcano plot representing differentially expressed proteins (DEPs) in *Oga*^K885N/+^ **(B)** and *Oga*^K885N/K885N^ **(C)** compared to wild type. The log2 fold changes are plotted against −log10(p-value), highlighting upregulated, downregulated, and non-significant DEPs. Significant proteins, including OGA are labelled. **(D)** Venn diagram representing the overlap of deregulated proteins in *Oga*^K885N/+^ and *Oga*^K885N/K885N^ compared to wild type mice. Deregulated proteins were defined as those with a *p* < 0.05. The fuchsia circle represents proteins deregulated in *Oga*^K885N/+^, the green circle represents those deregulated in *Oga*^K885N/K885N^. The intersection highlights the proteins that are commonly deregulated in both genotypes. **(E)** Cell compartment enrichment analysis of proteins commonly deregulated proteins in both *Oga*^K885N/+^ and *Oga*^K885N/K885N^ using Panther Classification system tool (94,95). The x-axis lists the enriched terms, ranked by fold enrichment. **(F)** Gene ontology (GO) enrichment analysis of proteins commonly deregulated proteins in both *Oga*^K885N/+^ and *Oga*^K885N/K885N^ using Metascape Analysis Resource tool (96). The x-axis lists the enriched terms, ranked by fold enrichment.

### Hippocampal Oga^K885N^ neurons show delayed maturation and disorganized synaptic activity

To investigate the effect of the K885N variant on synaptic activity and neuronal function, spontaneous neuronal spiking activity of primary hippocampal cultures from *Oga*^K885N^ mice was recorded at 7, 11, 14, and 17 days *in vitro* (DIV) using a High Density multi electrode array (HD-MEA) system. A summary of parameters measured and statistics is reported in **Supplemental Tables 1 and 2**. Hippocampal neurons from *Oga*^+/+^, *Oga*^K885N/+^ and *Oga*^K885N/K885N^ animals showed increased activity over time as seen by an increase in both spike amplitude and firing rate from DIV 7 to DIV 17 (**Fig. 7A-C**), corresponding to the development and maturation of excitatory synapses (48). In *Oga*^+/+^, spike amplitude increases rapidly and reached a plateau at DIV 14 (**Fig. 7B**) corresponding a peak of synaptogenesis (48). In contrast, both *Oga*^K885N/+^ and *Oga*^K885N/K885N^ neurons displayed slower rises of activity and significantly lower spike amplitudes compared to *Oga*^+/+^ hippocampal neurons at all recording days (**Fig. 7B**), suggesting a delay in synaptogenesis and synaptic maturation in *Oga*^K885N^ neurons. In addition, both *Oga*^K855N/+^ and *Oga*^K885N/K885N^ neurons showed an accelerated increase of the firing rate, coupled with a slower reduction of inter spike intervals (ISI) over time compared to wild type neurons (**Fig. 7C, D**). We then considered spike burst patterns (defined as a fast train of at least three spikes in the same channel with ISI inferior to < 200 ms), as they are considered more informative and reliable than single spikes (49,50), and are involved in plastic events (51,52). We observed that while the average burst duration remains stable over time for all three genotypes, both mutant *Oga*^K885N/+^ and *Oga*^K885N/K885N^ displayed longer bursts at all recording days compared to wild type (**Fig. 7E**). The percentage of channels that displayed at least one burst in the recording time rapidly increased from DIV 7 in *Oga*^+/+^ neurons to reach a plateau at DIV 11, whereas both *Oga*^K885N/+^ and *Oga*^K885N/K885N^ showed a more progressive increase of bursting events until DIV 17 (**Fig. 7F**). In addition, *Oga*^K885N/K885N^ neurons showed a higher number of isolated spikes outside burst events from DIV 11 compared to *Oga*^+/+^ and *Oga*^K885N/+^ neurons (**Fig. 7G**). Together with reduced bursting events, these data suggest that *Oga*^K885N^ neurons displayed disorganized and irregular spiking activity. In the hippocampus, LTP is associated with high-frequency post-synaptic firing, while LTD is associated with isolated spikes (53–55). Taken together, these data suggest that *Oga*^K885N^ neurons showed defects in synaptic plasticity, favouring LTD-like response.

**Figure 7:**
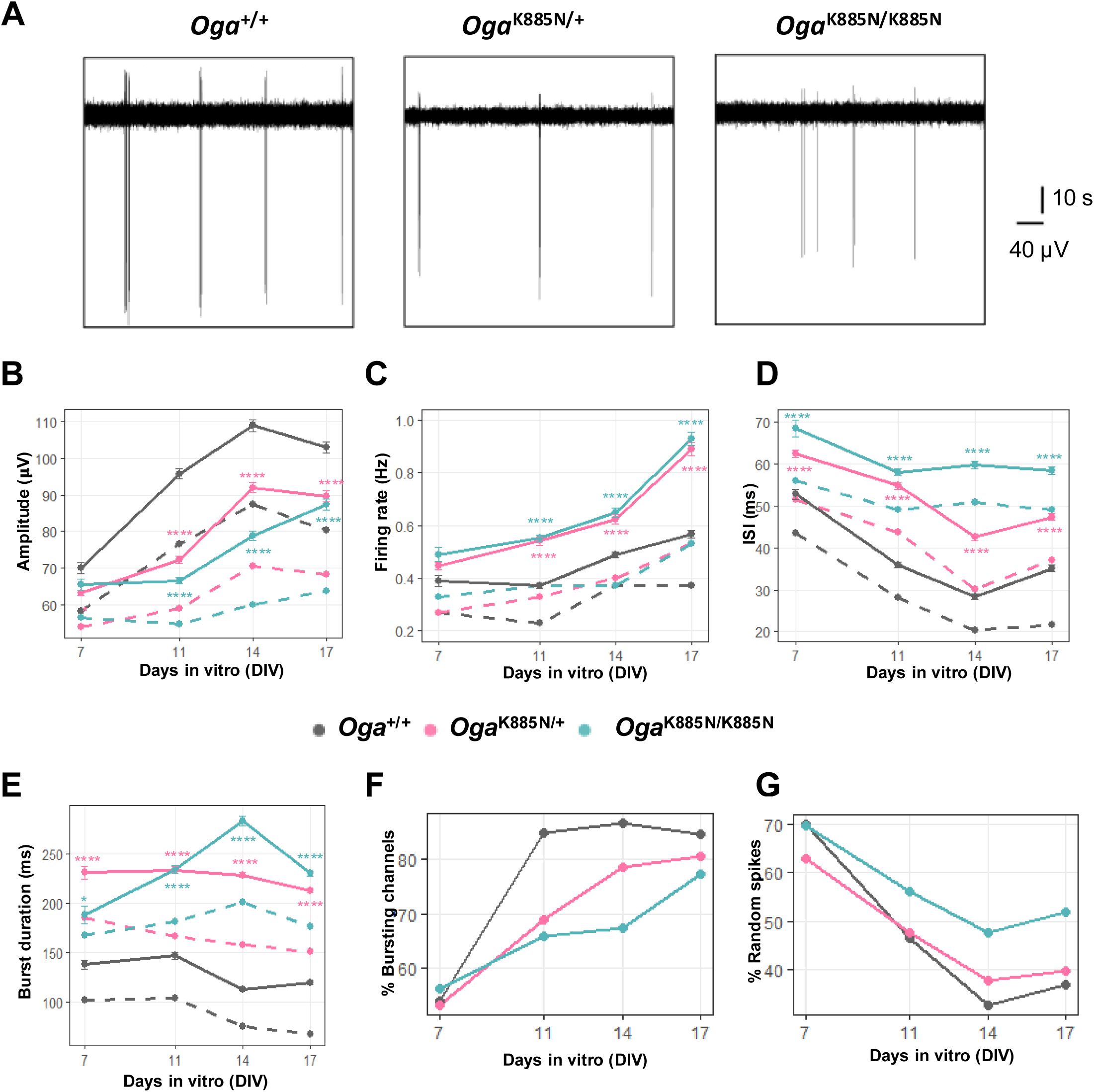
*Oga*^K885N^ mutation alters neuron spiking behaviour in primary hippocampal neurons. Data are represented both as means (solid lines) and medians (dashed lines). Significance using One-way Anova with Tukey multiple comparison is shown as * *p* < 0.05, ** *p* < 0.01 and *** *p* < 0.001. Number of replicates per genotypes are indicated in supplemental Table 6. **(A)** Representative neuron spontaneous activity traces of *Oga*^+/+^, *Oga*^K885N/+^ and *Oga*^K885N/K885N^ hippocampal neurons at DIV 17. Lower amplitude of spikes, lower channel burstiness, and higher spiking frequency can be observed in *Oga*^K885N^ hippocampal neurons compared to wildtype control. Measurement of spike amplitude **(B)**, firing rate **(C)** and inter spike interval (ISI) **(D)** of *Oga*^+/+^, *Oga*^K885N/+^ and *Oga*^K885N/K885N^ hippocampal neurons from DIV 7 to DIV 17. **(E)** Measurement of burst duration of *Oga*^+/+^, *Oga*^K885N/+^ and *Oga*^K885N/K885N^ hippocampal neurons from DIV 7 to DIV 17. **(F)** Percentage of channels that displayed at least one burst in *Oga*^+/+^, *Oga*^K885N/+^ and *Oga*^K885N/K885N^ hippocampal neurons from DIV 7 to DIV 17. **(G)** Percentage of spikes that do not belong to bursts in *Oga*^+/+^, *Oga*^K885N/+^ and *Oga*^K885N/K885N^ hippocampal neurons from DIV 7 to DIV 17.

## Discussion

Advances in genome sequencing have substantially accelerated the rate of identification of novel ID genes, revealing diverse molecular pathways critical for neurodevelopment. Pathogenic variants in *OGT* have previously been linked to ID (20), and functional studies of these variants consistently show secondary reductions in OGA protein levels, suggesting that perturbation of OGA may contribute to disease mechanisms (20,21,24,26,28–30). However, whether primary defects in OGA itself can cause human neurodevelopmental disease has remained unclear. Here, by integrating population genetics, clinical observations, and functional analyses, we provide converging evidence implicating OGA in human neurodevelopment and ID.

We report two unrelated individuals with ID carrying variants in *OGA*, representing the first such cases directly implicating OGA in ID. The clinical phenotypes differ in severity: the patient carrying the truncating OGA-Cys181* variant (and essentially heterozygous for *OGA*) presents with severe ID, epilepsy, dysmorphic features, generalized chorea, and microcephaly, whereas the patient carrying the OGA-K885N missense variant exhibits milder cognitive impairment with infrequent epilepsy and preserved brain structure.

To investigate the cellular and developmental consequences of *OGA* variation, we focused subsequent cellular and developmental analyses on the OGA-K885N variant, as the OGA-Cys181* nonsense variant is predicted to result in loss of the protein and to functionally resemble previously characterised heterozygous *Oga* knock-out models. Our data indicate that the OGA-K885N mutation reduces OGA protein levels through increased protein turnover, consistent with a posttranslational mechanism and resulting in effective heterozygosity. This reduction is independently observed in both *Oga*^K885N^ mESCs and mouse hippocampus tissues and, in contrast to both *Oga*^+/−^ mice and OGT-ID models (28–30,33), occurs without detectable changes in global O-GlcNAcylation. Despite preserved global O-GlcNAcylation levels, neurons carrying the K885N mutation exhibit impaired neuronal maturation and altered synaptic activity, while neuronal differentiation capacity and gross brain morphology remain intact.

Together, these findings indicate that partial reduction of OGA dosage can have measurable neurofunctional consequences that are uncoupled from global O-GlcNAc homeostasis. This dosage-sensitive effect is consistent with prior studies showing that complete or brain-specific loss of OGA in mice leads to severe developmental abnormalities, whereas heterozygous loss preserves gross brain structure (39,56). The distinct severity observed in the two individuals carrying *OGA* variants is therefore compatible with differing degrees of functional OGA impairment/levels.

Individuals with OGT-ID typically exhibit moderate to severe ID, dysmorphic features and brain anomalies. In these cases, reduced OGA protein levels have been reported as a compensatory response to impaired OGT activity and O-GlcNAcylation (20). The combined disruption of OGT function and secondary OGA downregulation may explain the severity of the OGT-ID variants. While it remains uncertain how the OGA-C181* variant affects O-GlcNAc homeostasis, the OGA-K885N variant leads to less than 50% reduction in OGA levels, insufficient to alter global O-GlcNAc levels as previously reported (9,11). This difference in molecular impact may contribute to the comparatively different clinical presentation.

Proteomics, behavioural and electrophysiological analyses further support a role for OGA dosage in neuronal circuit maturation and function. In particular, hippocampal neurons derived from Oga^K885N^ mice show temporal altered firing properties, including changes in spike frequency and amplitude, consistent with impaired synaptic maturation and neuronal excitability. Synaptic maturation is shaped by coordinated pre- and post-synaptic mechanisms and OGA has previously been implicated in the regulation of hippocampal synaptic morphology and plasticity. Particularly, OGA overexpression has been shown to promote synapse formation and stability through modulation of AMPA receptor subunit composition (57). Consistent with this, *Oga*^+/−^ mice display impaired hippocampal synaptic plasticity at SC-CA1 synapses, accompanied by dysregulated phosphorylation of AMPA receptor subunit GluA1 during chemically induced LTP and LTD (39). While our data indicate delayed synaptic maturation and disorganised network activity in *Oga*^K885N^ neurons, further work is required to elucidate whether these effects are mediated through AMPA receptor-dependent mechanisms or reflect broader disruptions of synaptic development as suggested by a broader scope of proteomic changes linked to synaptic structure and function. Importantly, electrophysiological alterations are accompanied by subtle behavioural changes, including increased compulsive or repetitive behaviours, while performance on assays probing hippocampus-dependent learning and memory remains largely preserved. This contrasts with reports from *Oga*^+/−^ mice, which display deficits in hippocampus-dependent learning and memory (39), suggesting that distinct neurofunctional consequences may arise depending on the extent and nature of OGA perturbation and its downstream effects. Together with reports that pharmacological inhibition of OGA produces distinct behavioural effects compared with reduced OGA expression (58), these findings support a model in which OGA protein dosage and enzymatic function exert partially dissociable effects on neuronal development, circuit maturation and behaviour.

The molecular mechanisms by which functional OGA levels affects brain development and function, however, remain to be fully elucidated. Notably, the K885N substitution is located in the C-terminal pseudo-histone acetyltransferase (pHAT) domain rather than the catalytic glycoside hydrolase domain. Together with the preserved global O-GlcNAcylation observed in *Oga*^K885N^ models, this points toward a potential contribution of non-catalytic OGA functions to the neurodevelopmental phenotypes. Human OGA exists in two main splice variants: a short isoform (OGA-S) that is expressed predominantly during prenatal brain development, and a long isoform (OGA-L) that is expressed throughout pre- and post-natal brain development in rodents (59). The OGA-L isoform is composed of an N-terminal glycoside hydrolase domain, a central stalk linker domain and a C-terminal pseudo-histone acetyltransferase (pHAT) domain, whereas the shorter isoform lacks the pHAT domain (60,61). While the catalytic domain mediates removal of O-GlcNAc from proteins, the biological role of the pHAT domain remains incompletely understood (62–64). Structural studies indicate that the pHAT domain adopts a fold related to GCN5 family acetyltransferases but lacks key residues required for acetyl-CoA binding and enzymatic activity (65). Notably, the OGA-K885N variant localizes to the pHAT domain, within a conserved region implicated in substrate or partner binding. Previous studies have linked OGA and its pHAT domain to transcriptional activation and regulation of chromatin states, including modulation of histone modifications such as H3K36 methylation and acetylation (42,43,66–68). Perturbation of this domain could therefore affect gene regulatory programs essential for neuronal maturation without altering global O-GlcNAc homeostasis, providing a plausible mechanistic framework for the phenotypes observed in *Oga*^K885N^ models that will require future investigation.

Our findings can be placed within a broader framework of convergent genetic architecture linking rare neurodevelopmental disorders to variation in cognitive function in the general population. In addition to identifying *de novo OGA* variants in individuals with intellectual disability, we show that both common non-coding variation and aggregated rare coding variation at the *OGA* locus associate with modest differences in cognitive performance in population-based datasets. This pattern mirrors that observed for established neurodevelopmental genes such as *SYNGAP1* (31,69,70), *CHD8* (71,72), *NRXN1* (72,73), and *KDM5B* (72,74), where rare, high-impact variants cause severe developmental phenotypes, while common alleles exert subtler effects on cognition and educational attainment. Together, these observations support a model in which OGA contributes to neurodevelopment and cognitive function across multiple layers of genetic variation.

Importantly, this dosage-sensitive role of OGA has implications beyond neurodevelopment. A substantial body of literature suggests that increasing O-GlcNAc levels through pharmacological inhibition of OGA may be beneficial in neurodegenerative disease contexts. However, most OGA inhibitors have failed in clinical trials, in part due to neurotoxicity (75). Together with our data, these findings suggest that perturbing OGA, either through reduced protein dosage or sustained enzymatic inhibition, can be deleterious for neuronal function. This underscores the need for caution when targeting O-GlcNAc cycling therapeutically and highlights the importance of preserving finely balanced OGA function for normal cognitive and neuronal processes.

In conclusion, we identify pathogenic variants in *OGA* in individuals with intellectual disability, providing the first direct evidence linking OGA to human neurodevelopmental disease. Functional analyses of the OGA-K885N variant demonstrate that reduced OGA protein dosage, despite preserved global O-GlcNAcylation, is sufficient to impair neuronal maturation and synaptic activity. Together, these findings establish OGA as a dosage-sensitive regulator of neurodevelopment and highlight the importance of tightly regulated OGA function for normal neuronal function.

## Materials & Methods

### Ethical permissions

The Comité de protection des personnes (CPP) ILE DE France VI (Genedefi - CPP 74-12) and the Bioethics Committee for Scientific Research of Medical University of Gdańsk (NKBBN/13/2023) gave ethical approval for this work. The participants’ parents provided written and informed consent.

### Fine-mapping of the MGEA5/OGA association signal in GWASs on cognition-related traits

Statistical fine-mapping was performed using the Genome-wide Complex Trait Bayesian analysis (GCTB) framework (76,77) with summary statistics from a published GWAS datasets of intelligence (36). Following fine-mapping, analyses were restricted to the *OGA* genomic region defined in GRCh37, corresponding to the genome assembly used in the source GWAS dataset. All variants reported in the summary statistics within the locus were included in the fine-mapping analyses without additional filtering. Variants were prioritised based on their posterior probabilities of causality, and most credible set of SNPs were visualized in a regional association plot capturing the broader linkage disequilibrium structure surrounding the association signal.

### Population-based analysis of OGA variation and cognition in the UK Biobank

A general cognitive ability (*g-score*) was derived using confirmatory factor analysis in *lavaan* (R v4.5.1), combining eight UK Biobank cognitive measures (fluid intelligence, matrix reasoning, numeric memory, symbol–digit substitution, trail-making, matching-task memory, and log-transformed reaction time), following the method as described in (78). In contrast to the original study, we did not incorporate socioeconomic indicators derived from UK census data.

We restricted the dataset to unrelated White British participants by excluding individuals with KING kinship coefficient ≥ 0.044, requiring self-reported British ethnicity (field 21000) and genetically inferred Caucasian ancestry (field 22006), removing samples with genetic missingness > 0.05 (field 22005), and excluding those with sex chromosome aneuploidy (field 2019). After quality control, 333,530 individuals remained.

To test whether common (MAF ≥ 1 %) variation at the *OGA* locus associates with general cognitive ability (g-score), we extracted all variants from Whole Genome Sequencing (WGS) data from the UK Biobank dataset DRAGEN pipeline within ±75 kb of the *OGA* gene chr10:101,709,443-101,893,465 (GRCh38). Variant-level quality control applied (Missingness < 2%, Hardy–Weinberg equilibrium p > 1×10⁻□:). To control for population structure, we used the imputed UK Biobank v3 dataset (HRC/UK10K+1000G reference) to calculate the principal components (PC1–PC20). Association testing was performed in PLINK 2.0 using an additive linear regression model:

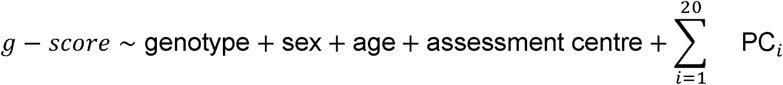

Residuals were inspected for normality, outliers, and heteroscedasticity. *P* values were corrected for the number of variants tested within the ± 75 kb locus.

All protein-altering variants (missense, protein-truncating, splice-region, and in-frame alleles) with MAF < 1 % and annotated to *OGA* were extracted from WGS data and annotated using Ensembl VEP, supplemented with CADD, ClinPred/Clin, and AlphaMissense pathogenicity scores. Because most alleles were extremely rare, we used per-variant carrier vs non-carrier comparisons rather than collapsing into a burden test. For each variant, we estimated:

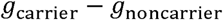

using covariate-adjusted linear models with the sex, age and assessment center as covariates. Given the heterogeneity and low frequency of individual alleles, estimates were not interpreted independently; emphasis was placed on aggregate patterns across all rare coding variants, including effect-size distributions and consistency with predicted pathogenicity. All analyses were performed under UK Biobank application 102638. All analyses were performed using PLINK2 (v2.00), PLINK1.9, R (v4.5.1), and Python (v3.14.0).

Each rare missense allele was mapped onto the canonical OGA protein structure (GH84 catalytic domain, stalk domain, pHAT domain). Domain-level variation was evaluated by combining CADD, Clin, and AlphaMissense scores to generate a positional pathogenicity profile, enabling assessment of structural constraint and depletion of variation in functionally important regions.

### Exome sequencing for OGA-ID variants

Library preparation was performed using SeqCap EZ MedExome kit (RocheTechnologies) and sequencing was generated on a NextSeq 500 instrument (Illumina Inc.) according to the manufacturer’s protocols. Both parents and the proband were analysed (trio exome sequencing). For data processing, raw reads were mapped to the human genome reference-build hg19 using the Burrows Wheeler Aligner (BWA MEM v0.717) alignment algorithm. The resulting binary alignment/map (BAM) files were further processed by Genome Analysis Tool Kit HaplotypeCaller (GATK HC v3.8). The VCF files were then annotated on Snpeff version 4.3T. Only coding non-synonymous and splicing variants were considered. Variant prioritization was conducted thanks to the transmission mode (*de novo*, autosomal recessive and X-linked), and the frequency of the variants in the GnomAD database.

### Transfection and Flow Cytometry Analysis

The dual fluorescently tagged OGT-sfGFP & mScarlet3-OGA mouse embryonic stem cells (mESCs) were cultured in 2i medium and transfected with plasmids using a previously described protocol (47). Briefly, 5.0 × 10c: cells were transfected with 1.5 µg of DNA plasmid and 3 µl of Lipofectamine 2000 (Thermo) for 24 h, according to the manufacturer’s instructions. After 48 h, cells were either directly analysed by flow cytometry to measure OGT-sfGFP and mScarlet3-OGA fluorescence or fixed for O-GlcNAc staining. For fixation, cells were incubated in 250c:µl of warm 4 % paraformaldehyde (PFA)/4 % sucrose for 10 min at room temperature. Subsequently, 1 ml of PBS was added, and the cells were centrifuged at 400 × g for 4 min. Cells were permeabilized in PBS buffer containing 5 % BSA and 0.3 % Triton X-100 for 1 h at room temperature. Cells were then centrifuged at 500 × g for 5 min and incubated overnight at 4c:°C in 100 µl of diluted blocking buffer (1 % BSA, 0.06 % Triton X-100 in PBS) containing 2 µl RL2-Alexa Fluor 647 antibody (Thermo Fisher). Cells were centrifuged at 500 × g for 5 min and resuspended in PBS for flow cytometry analysis.

Cells were analysed using a NovoCyte Quanteon 4025 flow cytometer (Agilent, Santa Clara, CA) equipped with four lasers (405 nm, 488 nm, 561 nm, and 637 nm) and 25 fluorescence detectors. Data acquisition was performed using NovoExpress software (v.1.6.2), and analysis was carried out using FlowJo (v.10). The gating strategy and flow cytometry detection channels for OGT-sfGFP, mScarlet3-OGA, mTagBFP2, and Alexa Fluor 647 were configured as previously described (42). Statistical analyses were performed using GraphPad Prism (v.10). An ordinary one-way ANOVA was used to compare groups.

### Cloning, protein expression and purification

A PCR product encoding OGA residues 11-396 was produced from a pre-existing full-length codon-optimised clone of human OGA obtained from Genscript. This fragment was subcloned as a *Bam*HI-*Not*I fragment into the plasmid pHEXmangled, a plasmid based on the pGEX6P1 backbone with the GST and PreScission sites replaced by a 6His tag. The same full-length template was used to produce a PCR product encoding the residues 536 to the end of human OGA preceded by a start codon. This was cloned as a *Xho*I-*Bam*HI fragment into pLinker2, a plasmid based on pET28a but lacking any encoded tag or start codon. The D175N and K885N mutations were introduced into plasmids Hm h133 3 and L2P LEND respectively to generate plasmids Hm A17 1 and L2P 85HE 4 respectively. The mutagenesis was carried out based on the Quikchange site directed mutagenesis kit by Stratagene, but using KOD polymerase instead of *Pfu*. All inserts were confirmed by DNA sequencing.

The OGA^WT^, OGA^K885N^ and catalytic defective OGA^D175N^ proteins were obtained by co-transforming a plasmid containing the N-terminal region of OGA (11–396) with another plasmid containing the C-terminal region of the protein (535-end) as previously described by (79). Briefly, OGA^K885^ was obtained by combining the Hm hi13 3 plasmid with the L2P LEND plasmid, OGA^K885N^ was obtained by combining the Hm hi13 3 plasmid with the L2P 85HE plasmid and finally, the OGA^D175N^ protein was obtained by combining the Hm A171 plasmid with the L2P LEND plasmid. A list of proteins and associated tags and constructs boundaries can be found in **Supplemental Table 3**. All constructs were transformed (or co-transformed, if needed) into *E. coli* BL21(DE3)pLysS for protein expression. Cell cultures were grown to an OD_600_ of 0.6 and induced with 300 mM IPTG at 16 °C overnight. The cultures were then harvested by centrifugation at 4 000 rpm for 20 min. Cells were resuspended in lysis buffer (25 mM Tris pH 7.5, 150 mM NaCl and 0.5 mM TCEP), supplemented with DNase, protease inhibitor cocktail (1 mM benzamidine, 0.2 mM PMSF and 5 µM leupeptin) and lysozyme and lysed using a French press device. Cell debris was removed by centrifugation, and the supernatant was incubated with Ni-NTA beads for 2 h on a roller at 4 °C. After an extensive wash of the beads with the lysis buffer, the recombinant proteins were cleaved off the beads with PreScission protease at 4 °C overnight. The cleaved proteins were then recovered from the beads, concentrated and loaded onto a 26/600 Superdex 200 column previously equilibrated with lysis buffer. Corresponding fractions were confirmed by SDS-PAGE, pooled, concentrated to 20 mg/ml, flash frozen in liquid nitrogen and stored at -80 °C until further use.

### Differential scanning fluorometry

Differential Scanning Fluorometry denaturing profiles were recorded with a BioRAD CFX Opus 96 Real-time PCR configured for detecting FRET signals. Briefly; 10 mM protein was mixed with 10 mM SYPRO Orange (Invitrogen ref S6650, 5X final working concentration) in 50 mM HEPES, 150 mM NaCl pH 7.5. Final reaction volume was 20 ml. Experiments were performed in sextuplicate, and results were plotted and further analysed using GraphPad Prism.

### Enzymology

Steady state kinetics of 4MU-GlcNAc (4-methylumbelliferyl-β-N-acetylglucosamine) were measured in buffer containing 25 mM HEPES (pH 7.5), with substrate concentrations ranging from 0 to 2 mM in three-fold dilutions. Reactions were performed with 2.5 nM enzyme in a 100 μl assay volume and at 37 °C. Product concentration (4-MU) was measured as fluorescence using a plate reader (Spectramax I3x) at an excitation wavelength of 365 nm and emission of 445 nm. Experiments were performed in triplicate, and Michaelis-Menten kinetics were analysed using GraphPad Prism.

### Generation of mESC cell lines carrying OGA^K885N^ variant

AW2 mESCs derived from E14-TG2a.IV (129/Ola) ES cells were kindly donated from the MRC Centre for Regenerative Medicine, Institute for Stem Cell Research, University of Edinburgh (80) and cultured as previously described (21). Editing CRISPR reagents including pBABED and pX335 were prepared as previously described (81). Mouse embryonic stem cells were transfected using Lipofectamine 3000 (Invitrogen, according to manufacturer’s instructions) with pBABED, pX335 and the repair templates coding for either the K885N mutation or the native lysine, to obtain *Oga*^+/+^, *Oga*^K885N/+^ and *Oga*^K885N/K885N^ clones. After 24 h, transfected cells were selected using 1 μg/ml puromycin treatment for 2 days. Colonies from single cells that survived puromycin treatment were grown for 1-2 weeks, genotyped and sequenced to confirm the insertion of repair templates. Editing CRISPR reagents and primers sequences are listed in **Supplemental Table 4**. For subsequent experiments, mESCs were cultured in 2i media.

### Whole-genome sequencing and variant analysis of genome edited mESCs

Whole-genome sequencing (WGS) was performed on genome-edited mESC clones to confirm correct insertion of the K885N allele and to exclude off-target editing events. High-molecular-weight genomic DNA was extracted using DNeasy Blood and Tissue Kit (Quiagen) and prepared for sequencing following manufacturer protocols. Paired-end libraries were sequenced on an Illumina platform to a mean coverage of approximately 30×. Data were processed using nf-core/sarek v3.4.2 (82–84) of the nf-core collection of workflows (85), utilising reproducible software environments from the Bioconda (86) and Biocontainers projects. The pipeline was executed with Nextflow v24.04.4. (87) Specifically, FASTQ files were pre-processed with Fastp and aligned to the mouse reference genome (GRCm38/mm10) using BWA-MEM and resulting BAM files were processed following the GATK Best Practices workflow, including duplicate marking, indel realignment, and base quality recalibration.

Variant calling was performed with GATK HaplotypeCaller, generating per-sample gVCFs subsequently joint-genotyped across all edited clones. Annotation of variants was performed using SnpEff. A multi-step filtering strategy was applied to identify potential off-target edits: 1) Removal of known background variation, 2) Exclusion of variants shared across all clones, and 3) Hard-filtering of remaining variants following GATK recommendations: ‘QD >= 2 && FS <= 60 && SOR <= 3 && MQ >= 40’. (https://gatk.broadinstitute.org/hc/en-us/articles/360035890471-Hard-filtering-germline-short-variants) From the remaining variants, only missense and nonsense variants were kept. Finally, to distinguish true editing-induced changes from background variation, we retained only variants that were present in *Oga*-targeted clones, but consistently homozygous reference in a set of in-house control clones generated from the same parental mESC line with CRISPR edits at unrelated loci.

Candidate sites within or proximal to predicted CRISPR off-target regions were manually reviewed in IGV, and the *Oga* locus was inspected to verify correct insertion of the K885N allele. Only clones harbouring the precise K885N variant and showing no additional coding variants in *Oga* and nearby regions were selected for downstream cellular, biochemical, and differentiation experiments.

### Cycloheximide assay

mESCs cells were treated with cycloheximide (Cayman Chemical) at a final concentration of 20 μg/ml for 0, 2, 6, 10 and 24 h before lysis. DMSO was used as a control.

### Neuronal differentiation of mESCs

mESCs were differentiated as described (88). Briefly, cells were plated into a non-adhesive bacterial dish in basal differentiation media I (DMEM/F12, 10 % FBS, 1 % NEAA and 0.1 mM 2ME) for 8 days to induce embryoid body formation. From day 4 to day 8, cells were treated with 1 µM retinoic acid (R2625; Sigma-Aldrich) to obtain NPCs. At day 8, cells were plated on glass slide or petri dishes coated with poly-D-lysine and mouse laminin in N2 media (DMEM/F12 and 1 % N2). At day 10, media was changed into N2B27 media (50 % DMEM/F12, 50 % Neurobasal media,1 % N2, 1 % B27). Cells were harvested at day 18 for further analysis.

### Western blots

Proteins were extracted in RIPA lysis buffer (Cell signaling) for 30 min on ice, centrifuged at 14,000 rpm for 20 min at 4 °C and protein concentration was determined with Pierce 660 nm protein assay (Thermo Scientific). Proteins were then separated on precast 4 to 12 % NuPAGE Bis–Tris Acrylamide gels (Invitrogen) and transferred to a nitrocellulose membrane. Membranes were incubated with primary antibodies overnight at 4 °C or 3 h at room temperature. Anti-OGA (1:500 dilution; HPA036141; Sigma), anti-O-GlcNAc (RL2) (1:500 dilution; NB300-524, Novus Biologicals), anti-OGT (F-12) (1:1000 dilution; sc-74546; Santa Cruz), rabbit anti-C2orf70/FAM166C (1:1000 dilution; NBP2-82897; Novus Biologicals), rabbit anti-actin (1:2000 dilution; A2103; Sigma), mouse anti-lamin B (1:10000 dilution; 66095-Ig; Proteintech) and rabbit anti-lamin B (1:5000; 12987-1-AP; Proteintech). IR680/800-labelled secondary antibodies (Licor) were used for detection. Blots were imaged using a DLx Odyssey infrared imaging system (Li-Cor), and signals were quantified using Image Studio Lite (Li-Cor) or Empiria Studio (Li-Cor). Results were normalized to the mean of each corresponding wild type (*Oga*^+/+^) replicates set and represented as a fold change relative to *Oga*^+/+^.

### qPCR analysis

Total RNA was purified from cells using RNAeasy Kit (Qiagen), and then 0.5-1 µg of sample RNA was used for reverse transcription with the qScript cDNA Synthesis Kit (Quantabio). Quantitative PCR reactions were performed as previously described (11). Samples were assayed in biological replicates with technical triplicates using a BioRAD CFX Opus 384 Real-time PCR. Results were normalized to the mean of each corresponding *Oga*^+/+^ replicate set and represented as a fold change relative to *Oga*^+/+^. A list of primers used can be found in **Supplemental Table 2**.

### Immunostaining

Cells were fixed in 4 % PFA for 15 min at room temperature, followed by permeabilisation with 0.3 % Triton-X for 10 minutes. Blocking was performed for 1 h at room temperature with 5% of donkey serum in PBS. Primary antibody for Sox2 (1:200; #23064; Cell Signaling Technology), Nanog (1:200; #8822; Cell Signaling Technology), NeuN (1:200; ab104224; Abcam) and βIII tubulin (1:500; MAB1195; R&D Systems) were prepared in blocking solution and incubated overnight at 4°C. Secondary antibodies (1:1000; Alexa Fluor™ 488 A-21202 and Alexa Fluor™ 568 A10042; ThermoFisher) were prepared in blocking buffer and incubate for 2 h at room temperature in the dark. DAPI (62248; ThermoFisher) was prepared in PBS and incubate for 5 min at room temperature. Each incubation was followed by 3 washes in PBS. Dako media was used for mounting.

### Generation of the Oga^K885N^ mice and animal husbandry

*Oga*^K885N^ mice were produced by microinjection under project licence PPL PB0DC8431 at the Central Transgenic Core of Bioresearch and Veterinary Services at the University of Edinburgh, UK as previously described (28). Editing CRISR reagents, Cas9 nickase (1081062; Integrated DNA Technologies) and primers were purchased from Integrated DNA Technologies, and sequences are listed in **Supplemental Table 5.** Genomic DNA from offspring was genotyped and sequenced to confirm the presence of the *Oga*^K885N^ variant. The *Oga*^K885N^ line was maintained under C57BL/6J background (Janvier, France) for further breeding. Only male mice were used in all experiments. Animals were housed in ventilated cages with water and food available ad libitum and 12/12 h light/dark cycles in the Skou animal facility of Aarhus University. All animal studies and breeding were performed in accordance with the ARRIVE guidelines and the European Communities Council Directive (2010/EU) and were approved by the Danish Animal Experiments Inspectorate (Dyreforsøgstilsynet), under Breeding license 2022-15-0202-00135 and Project licenses: 2023-15-0201-01426.

### Behaviour

All behaviour experiments were performed as previously described (29). Animal cohorts were obtained from crossing of heterozygous male *Oga*^K885N/+^ with heterozygous female *Oga*^K885N/+^. Only male *Oga*^+/+^ (n = 14), *Oga*^K885N/+^ (n = 17) and *Oga*^K885N/K885N^ (n = 15) were used for all the tests. The Dixon’s Q test was performed for outlier identification in the marble test for *Oga*^K885N/+^ group according to Rorabacher (89). The experimental Q value (Q_exp_) was calculated from raw data using Q_exp_ = (x_2_ − x_1_)/(x_n_ − x_1_), where x_1_ is the value of interest, x_2_, the closest value to x_1_ in the data set, and x_n_ the highest value in the data set. Q_exp_ was next compared with the Q critical value (Q_crit_) of 0.365 for n = 17 for a confidence level of 95 % using the Q-table. Data that displayed a Q_exp_ higher than the Q_crit_ of 0.365 were considered as an outlier at a confidence level of 95 %. Behaviour experiment and analysis were performed blind to genotypes.

### Mass spectrometry analysis

Proteins (100 µg) extracted from cortex and hippocampus isolated from *Oga*^+/+^, *Oga*^K885N/+^ and *Oga*^K885N/K885N^ (n = 4 per genotype) were prepared using a S-Trap™ 96-well Mini Plate (Protifi) according to the manufacturer’s recommendation, including three washes with 50 % CHCl₃/50 % MeOH. Samples were digested using sequence grade trypsin (Proteomics grade, Sigma-Aldrich) with a 1:50 ratio (w/w). The tryptic peptides were lyophilized and dissolved in 0.2% formic acid. LC-MS/MS was conducted using an EASY-nLC 1200 system (Thermo Scientific) connected to an Orbitrap Eclipse Tribrid Mass Spectrometer (Thermo Scientific) with a 2 cm trap column (100 μm i.d.) and a 20 cm analytical column (75 μm i.d.), both packed in-house with ReproSil-Pur C18-AQ 3 μm resin (Dr. Maisch GmbH). Peptides were eluted at 250 nl/min using a 115-minute gradient from 5 % to 44 % phase B (0.1 % formic acid and 80 % acetonitrile), followed by 100 % phase B for 5 minutes. Protein identification and quantification were performed using Proteome Discoverer 2.5 (Thermo Scientific). Data were searched against the mouse reference proteome (uniprot.org) using the Sequest search engine with the following parameters: MS error tolerance of 10 ppm, MS/MS error tolerance of 0.02 Da, trypsin as the protease with two missed cleavages, and carbamidomethylation as a fixed modification. Label-free quantification was based on precursor ions quantified in at least 70 % of the replicates. Peptide intensities were normalized to total peptide intensity and scaled using the average of all samples. Protein ratios were based on summed peptide abundances with imputation using low abundance resampling. Significantly regulated proteins were identified using ANOVA, with adjustments for multiple testing.

### Primary hippocampal cultures and HD-MEA analysis

Individual cultures of mixed neuronal and glial cells were prepared from postnatal days 1-3 newborn male pups from the Oga^K855N^ line. Hippocampi were dissected in ice-cold Dulbecco’s PBS (Gibco), dissociated by pipetting using TrypLE (Gibco) and incubated for 20 min at 37 °C. The digestion reaction was stopped by adding DMEM supplemented with 10% FBS, followed by centrifugation at 1200 *g* for 5 min. The cell pellets were resuspended in filtered Neurobasal medium [Neurobasal (Gibco) supplemented with 0.5 mM GlutaMAX, 2 % B-27 supplement (Gibco), and 1 % Penicillin-Streptomycin (Gibco). Single replicate Neuronal spiking activity was recorded using a 6-well, high-density multi-electrode array (HD-MEA) MaxTwo system (MaxWell Biosystems). HD-MEA plates were sterilized and prepared according to the manufacturer’s instructions. Two days before cell plating, plates were incubated with complete Neurobasal medium at 37 °C and 5 % CO_2_ and coated with 100 µg/ml Poly-L-lysine hydrobromide (Sigma-Aldrich) in Borate Buffer (Thermo Fisher) overnight at 37 °C and 5 % CO_2_. Approximately 80,000 dissociated hippocampal cells from single pup in 50 µl of medium were seeded on the active sensing area of the MaxTwo plates and incubated at 37 °C and 5 % CO_2_ for 1 h. After cell adhesion to the surface of the chip, 1.2 ml of complete Neurobasal medium was added to each well, and half of the medium volume was changed every 2-3 days. Cell culture, MEA recording and analysis were performed blindly.

Neuron spiking activity was recorded on days *in vitro* (DIV) 7, 11, 14 and 17 using the Activity Scan protocol in the MaxTwo software (MaxWell Biosystems) by recording every electrode on the chip for 30 seconds at 10 kHz and 512× gain. Data were filtered through a 300 Hz high-pass filter, and the spike detection threshold was 5.5 standard deviations. Only active electrodes (average spike amplitude > 20 µV, firing rate > 0.13 Hz) were used in the analyses. The full dataset was imported to and visualized in R. To account for the difference in animals used from each genotype, the total N of active electrodes was randomly downsampled to a maximum of 2000 per combination of genotype and DIV (**Supplemental Table 6**).

Distribution of average spiking parameters at a channel level were fitted in a mixed effects model. Distributions of the logarithm of average channel spike amplitude (Eq. 1) and of the logarithm of average channel inter spike interval within bursts (ISI, Eq. 3) were fitted in a linear mixed-effects model with a polynomial DIV component to represent the non-linear changes of these parameters over days in culture. Average channel firing rate was fitted into a gamma generalized mixed-effect model with a log link (Eq. 2), with a polynomial DIV component. A hierarchical random effect plate-well-electrode was included into each model to account for experimental and equipment-related variability. The models were estimated in R using the packages lme4 (90), and glmmTMB (91).

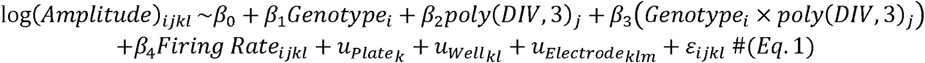

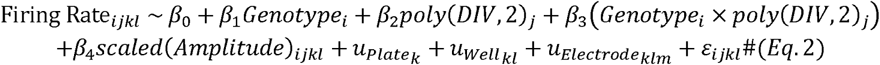

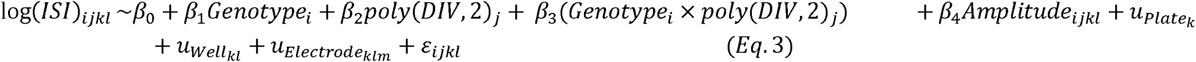

## Data Availability

All data produced in the present study are available upon reasonable request to the authors

## Acknowledgements

This work was funded by a Wellcome Trust Investigator Award (110061), a Novo Nordisk Foundation Laureate award (NNF21OC0065969) and a Villum Fonden Investigator (00054496) to D.M.F.v.A. Access to UK Biobank data was granted under Application ID 102638, which supported the analyses presented in this study. We would like to thank Bartosz Kowalski for support with enzymatic assay and Conor W. Mitchell for support with cycloheximide assay.

## Author contributions

F.A. and D.M.F.v.A conceived the study; F.A., B.A., O.S.Q., K.S.C., S.G.B., H.Y., I.E.A and C.S. performed experiments; A.T.F. performed molecular biology; D.D, P.C., C.M., M.K and M.M.B. collected clinical data; B.K. and P.Q. performed genomic analysis; F.A., B.A., O.S.Q., K.S.C., S.G.B., H.Y., I.E.A., M.P.V., C.S., P.D.R., P.Q. and D.M.F.v.A analysed data and F.A. and D.M.F.v.A interpreted the data and wrote the manuscript with input from all authors.

## Supplementary figure legends

**Supplementary Figure S1:**
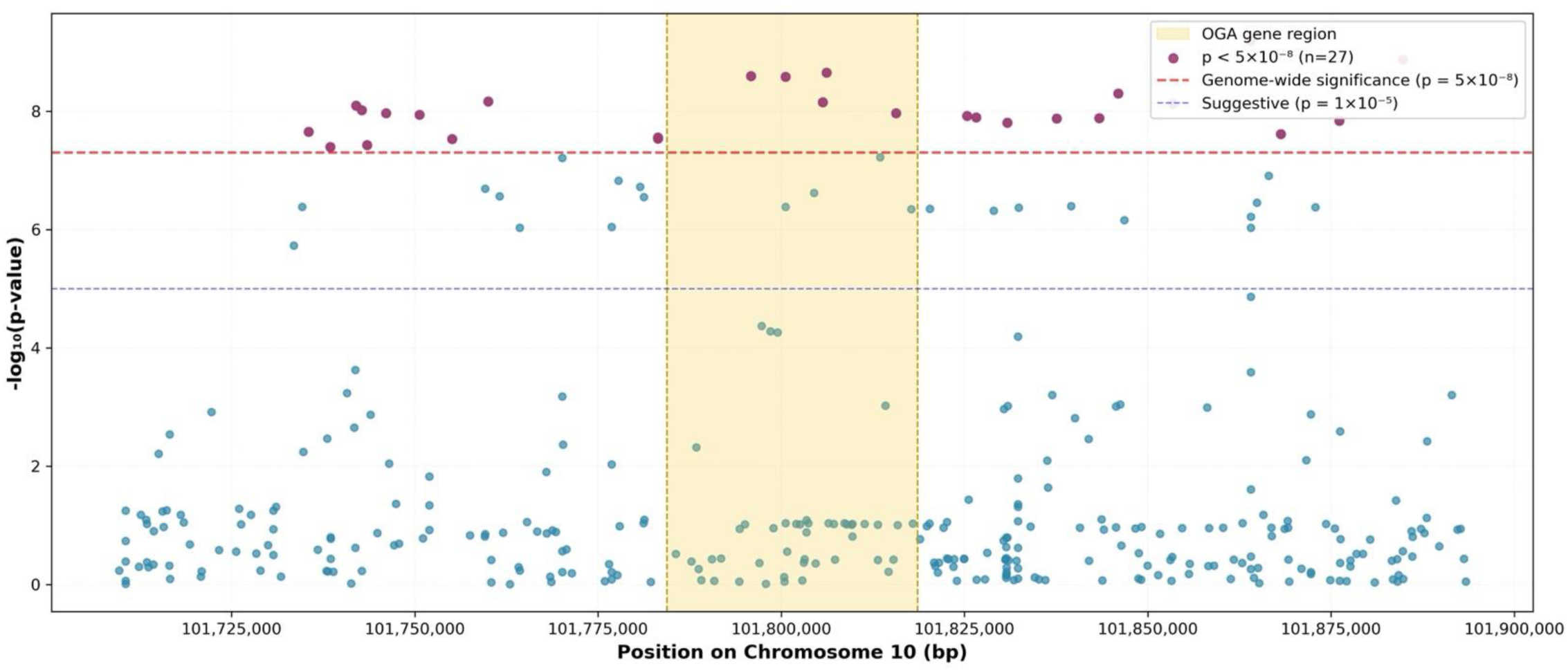
Manhattan plot of association signals for 337 common genetic variants (MAF ≥ 1 %) across the extended *OGA* locus (101,709,443–101,893,620 bp, GRCh38; ±75 kb flanking the gene boundaries). The *OGA* gene region (101,784,443–101,818,620) is shown in light yellow. The y-axis shows −log₁₀(p-values) from linear regression adjusted for sex, age, assessment center, and 20 genome-wide principal components, with variants plotted along the x-axis by genomic position. Genome-wide significant associations (p < 5 × 10⁻c:; red dashed line) are highlighted in purple, and a secondary threshold for suggestive associations (p < 1 × 10⁻c:; blue dashed line) is also shown. Analyses included 333,530 unrelated UK Biobank participants of British Caucasian ancestry with available g-score measurements and quality-controlled genotype data.

**Supplementary Figure S2:**
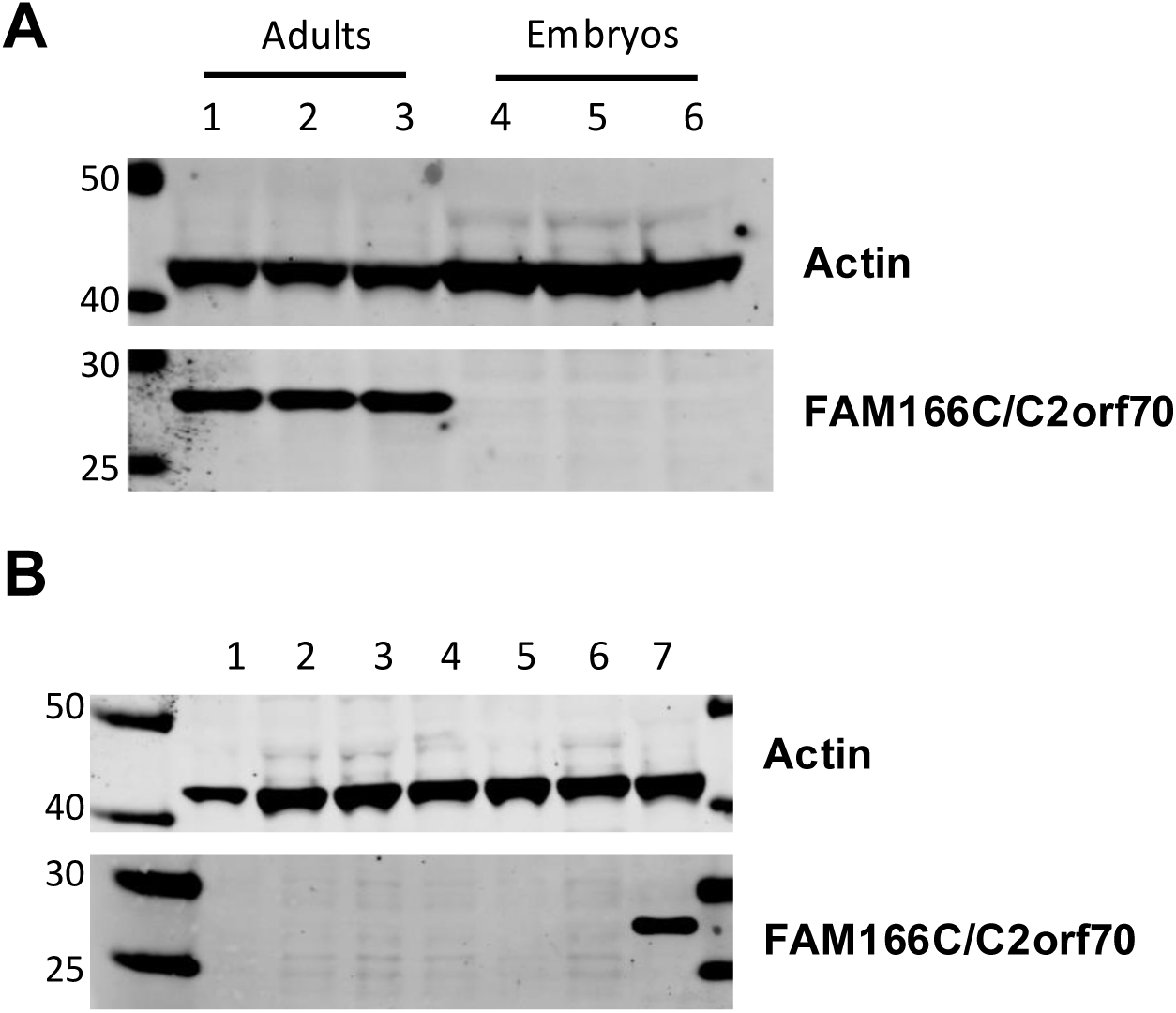
**(A)** Western blot of C2orf70/FAM166C protein levels in brain tissues from wild-type adult and E15.5 embryos mice. Actin antibody was used as loading control. **(B)** Western blot of C2orf70/FAM166C protein levels in wild-type undifferentiated mESCs (sample from 1 to 6) and brain from wild type adult mouse as control (sample 7). Actin antibody was used as loading control.

**Supplementary Figure S3:**
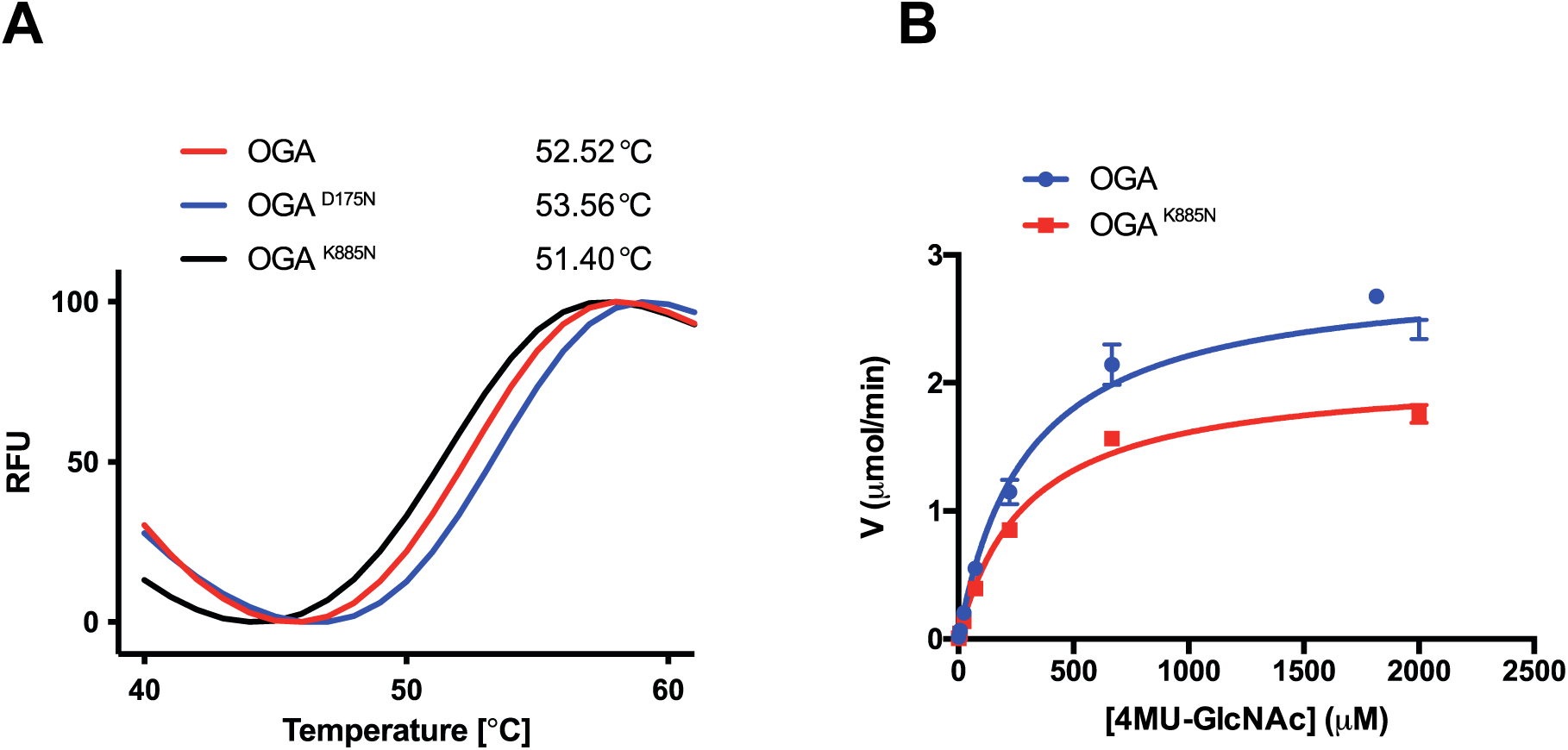
Effect of the K885N variant on OGA protein stability and catalytic activity. **(A)** Thermal denaturation profiles of OGA, OGA^K885N^ and OGA^D175N^ proteins. The *T_m_* values from OGA (52.5 ± 0.4 °C), OGA^K885N^ (53.6 ± 0.5 °C) and OGA^D175N^ (51.4 ± 0.5 °C) were calculated as the fluorescence vs. the temperature function first derivative maximum peak. Each graph data point represents the averaged data from 6 replicates. **(B)** Michaelis-Menten kinetics of the OGA and OGA^K885N^ constructs. The OGA kinetic values are, *V*_max_ = 2.9 ± 0.1 µM min^−1^ and *K*_M_ = 3.0 ± 0.3 µM while the OGA^K885N^ kinetic values are, *V*_max_ = 2.1 ± 0.1 µM min^−1^ and *K*_M_ = 2.9 ± 0.2 µM. Averaged data of 3 replicates is represented in the graph.

**Supplementary Figure S4:**
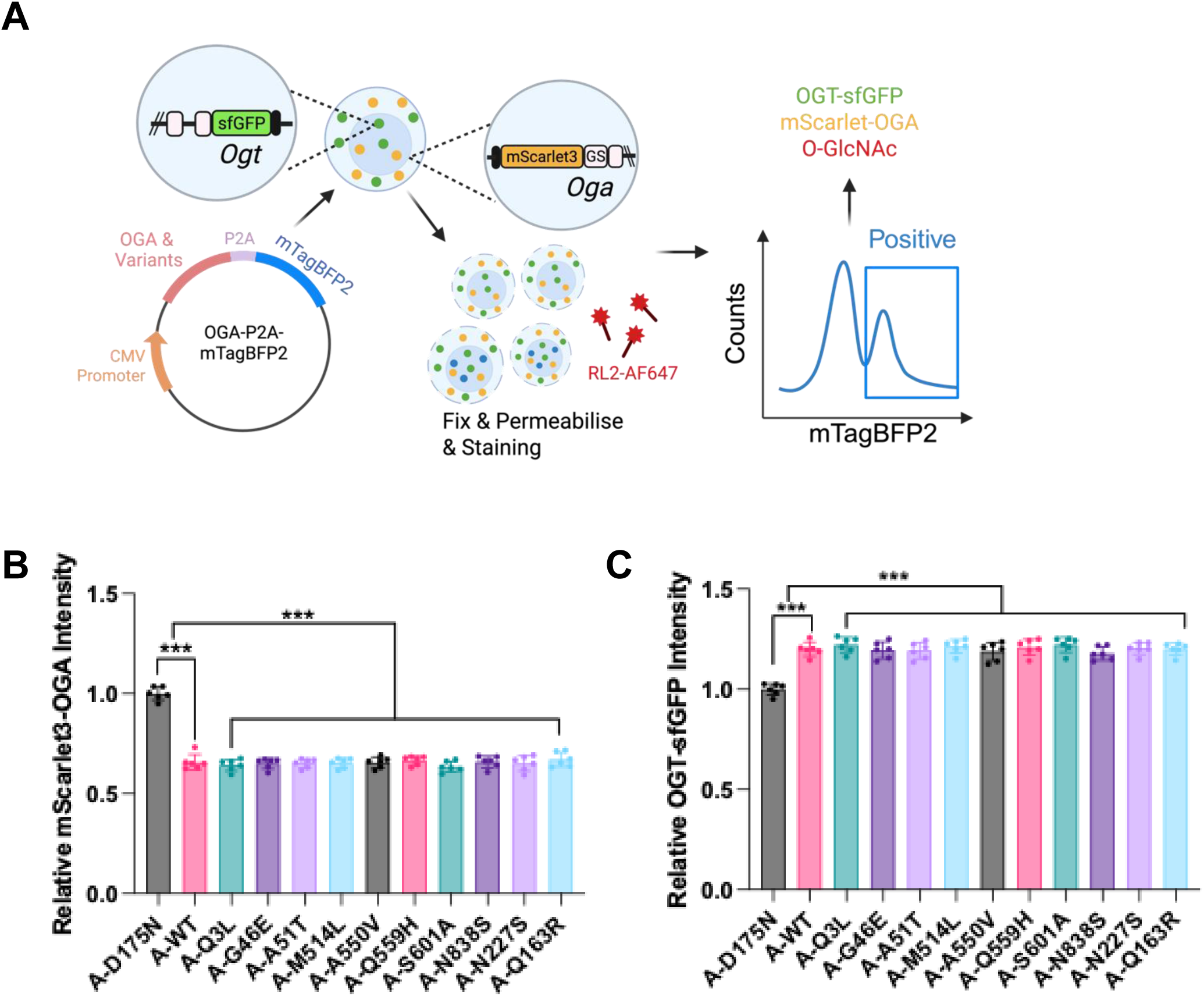
Effects of common (non-pathogenic) OGA variants on O-GlcNAc homeostasis. **(B)** Schematic overview of assessing OGA variants using the dual-tagged fluorescent system. Human OGA and its variants were cloned with a P2A self-cleaving sequence followed by the fluorescent marker mTagBFP2 and transfected into the previously described dual-tagged OGT-sfGFP & mScarlet3-OGA mouse embryonic stem cells (mESCs) (44). 48 h post-transfection, cells were either fixed and stained with the O-GlcNAc antibody RL2 conjugated to Alexa Fluor 647 or left unstained. Transfected (mTagBFP2-positive) cells were analysed for endogenous OGT, OGA, and, where applicable, O-GlcNAc levels. Endogenous levels of mScarlet3-OGA **(B)** and OGT-sfGFP **(C)** in dual-tagged mESCs measured by FACS after transfection with a selected panel of common OGA variants from the gnomAD database, representing variants distributed across each major domain of the OGA protein. The catalytically inactive OGA^D175N^ mutant served as a control and baseline for normalising all signals. Endogenous levels of mScarlet3-OGA.

**Supplementary Figure S5:**
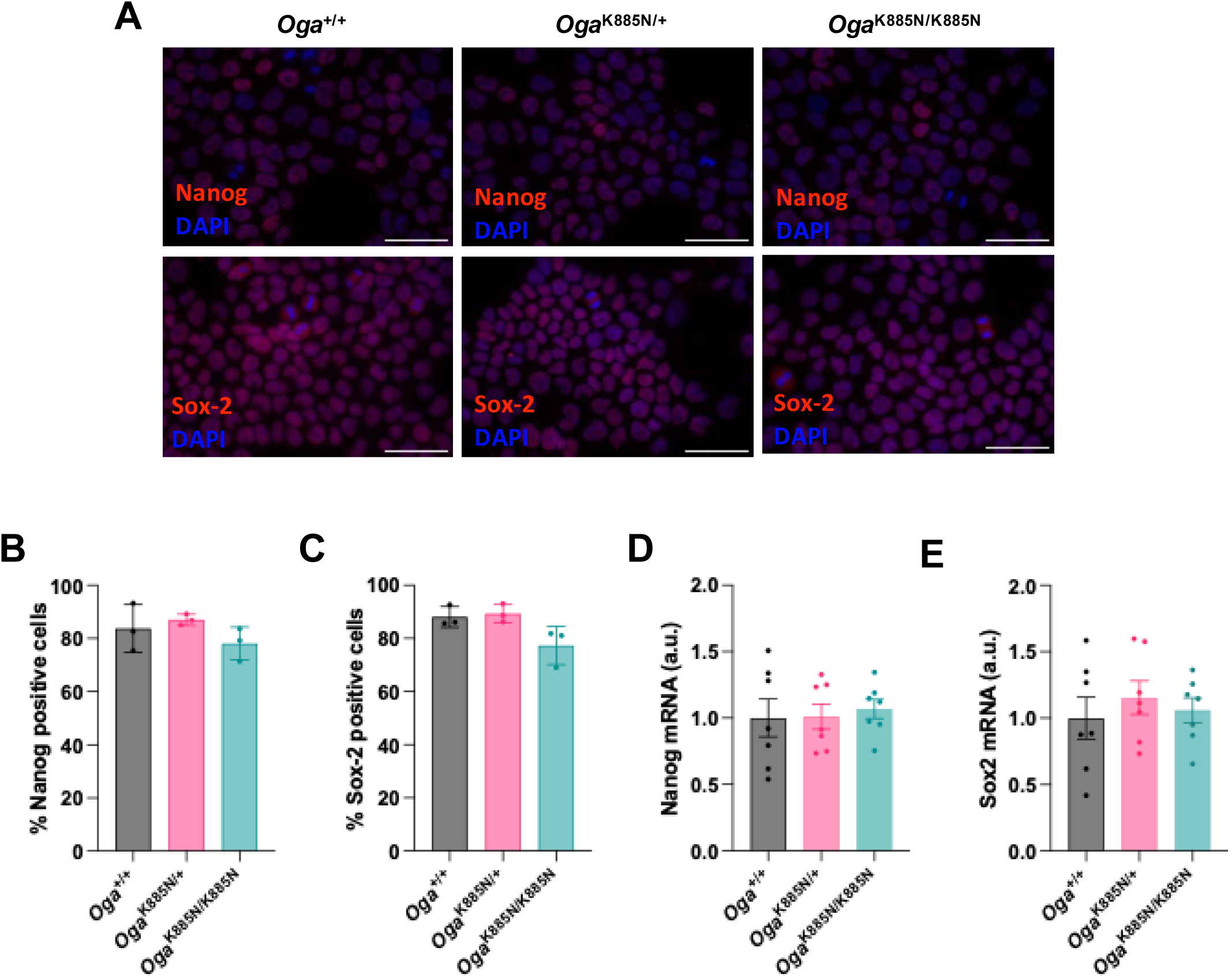
The K885N mutation does not affect pluripotency or self-renewal. Data are represented as mean ± SD, of minimum n = 3 independent experiments for all genotypes. Significance using One-way Anova with Tukey multiple comparison is shown as * *p* < 0.05 and ** *p* < 0.01**. (A)** Representative fluorescent microscopy images of Nanog and Sox2 expression in *Oga*^+/+^, *Oga*^K885N/+^ and *Oga*^K885N/K885N^ mESCs cells in undifferentiated conditions. Magnification 63x, 50 μm scale bar is indicated. Number of Nanog **(B)** and Sox2 **(C)** positive cells from panel A. Quantification of *Nanog* **(D)** and *Sox-2* **(E)** mRNA levels in *Oga*^+/+^, *Oga*^K885N/+^ and *Oga*^K885N/K885N^ mESCs cells in undifferentiated conditions.

**Supplementary Table 1:** Summary of values parameters for the most relevant spiking parameters measured using HD-MEA. Amplitude and firing rate are calculated per channel, while burst duration and spikes per burst are calculated per burst.

| Genotype | DIV | Amplitude mean (μV) | Amplitude median (μV) | Amplitude q25 (μV) | Amplitude q75 (μV) | Firing rate mean (Hz) | Firing rate median (Hz) | Firing rate q25 (Hz) | Firing rate q75 (Hz) |
| --- | --- | --- | --- | --- | --- | --- | --- | --- | --- |
| *Oga*<sup>+/+</sup> | 7 | 70.05 | 58.26 | 47.26 | 76.49 | 0.39 | 0.27 | 0.17 | 0.47 |
|  | 11 | 95.78 | 76.45 | 55.59 | 115.85 | 0.37 | 0.23 | 0.17 | 0.43 |
|  | 14 | 109.00 | 87.36 | 60.52 | 133.48 | 0.49 | 0.37 | 0.23 | 0.63 |
|  | 17 | 102.93 | 80.32 | 56.55 | 126.24 | 0.57 | 0.37 | 0.20 | 0.70 |
| *Oga*<sup>K885N/+</sup> | 7 | 63.17 | 53.79 | 44.27 | 71.11 | 0.45 | 0.27 | 0.17 | 0.50 |
|  | 11 | 72.24 | 58.81 | 46.95 | 82.00 | 0.54 | 0.33 | 0.20 | 0.60 |
|  | 14 | 92.00 | 70.53 | 50.69 | 111.39 | 0.62 | 0.40 | 0.23 | 0.73 |
|  | 17 | 89.75 | 68.29 | 49.98 | 104.23 | 0.89 | 0.53 | 0.23 | 1.10 |
| *Oga*<sup>K885N/K885N</sup> | 7 | 65.50 | 56.36 | 46.09 | 72.68 | 0.49 | 0.33 | 0.20 | 0.63 |
|  | 11 | 66.58 | 54.64 | 43.58 | 75.06 | 0.55 | 0.37 | 0.20 | 0.66 |
|  | 14 | 78.77 | 59.91 | 44.07 | 90.55 | 0.65 | 0.37 | 0.20 | 0.76 |
|  | 17 | 87.33 | 63.81 | 45.78 | 103.42 | 0.99 | 0.53 | 0.27 | 1.20 |
| Genotype | DIV | Burst duration mean (ms) | Burst duration median (ms) | Burst duration q25 (ms) | Burst duration q75 (ms) | Spikes per burst mean | Spikes per burst median | Spikes per burst q25 | Spikes per burst q75 |
| *Oga*<sup>+/+</sup> | 7 | 137.77 | 101.50 | 67.63 | 177.78 | 4.19 | 3 | 3 | 5 |
|  | 11 | 146.68 | 103.90 | 58.30 | 177.10 | 4.86 | 4 | 3 | 5 |
|  | 14 | 112.73 | 74.80 | 25.70 | 140.80 | 4.63 | 4 | 3 | 5 |
|  | 17 | 120.67 | 67.10 | 30.30 | 136.35 | 4.74 | 4 | 3 | 5 |
| *Oga*<sup>K885N/+</sup> | 7 | 231.43 | 185.05 | 94.13 | 285.28 | 4.52 | 4 | 3 | 5 |
|  | 11 | 254.54 | 166.85 | 92.00 | 273.40 | 5.28 | 4 | 3 | 5 |
|  | 14 | 238.06 | 157.80 | 75.70 | 277.63 | 5.38 | 4 | 3 | 6 |
|  | 17 | 219.54 | 151.10 | 69.30 | 262.25 | 5.28 | 4 | 3 | 6 |
| *Oga*<sup>K885N/K885N</sup> | 7 | 188.24 | 167.40 | 99.50 | 241.48 | 3.73 | 3 | 3 | 4 |
|  | 11 | 237.95 | 181.40 | 99.60 | 287.35 | 4.66 | 4 | 3 | 5 |
|  | 14 | 289.51 | 201.20 | 111.95 | 328.10 | 5.25 | 4 | 3 | 5 |
|  | 17 | 231.45 | 176.80 | 93.00 | 284.30 | 4.80 | 4 | 3 | 5 |

**Supplementary Table 2:** Estimators from the linear mixed-effects models used to determine the effect of genotype on different spiking parameters measured using HD-MEA.

| log(Amplitude) ~ Genotype × DIV(polynomial, 3) + Firing rate |  |  |  |  |
| --- | --- | --- | --- | --- |
|  | Estimate (β) | S.E. | t-value | p |
| Intercept | 3.9914 | 0.3697 | 10.796 | < 0.001 |
| *Oga*<sup>K885N/+</sup> | 1.7907 | 0.5011 | 3.574 | < 0.001 |
| *Oga*<sup>K885N/K885N</sup> | 1.6798 | 0.5140 | 3.268 | < 0.01 |
| DIV<sup>1</sup> | -0.1488 | 0.0979 | -1.520 | ns |
| DIV<sup>2</sup> | 0.0223 | 0.0084 | 2.651 | < 0.01 |
| DIV<sup>3</sup> | -0.0007 | 0.0002 | -3.217 | < 0.01 |
| Firing rate | 0.1575 | 0.0044 | 35.967 | < 0.001 |
| *Oga*<sup>K885N/+</sup> × DIV<sup>1</sup> | -0.4193 | 0.1370 | -3.060 | < 0.01 |
| *Oga*<sup>K885N/K885N</sup> × DIV<sup>1</sup> | -0.3478 | 0.1366 | -2.546 | < 0.05 |
| *Oga*<sup>K885N/+</sup> × DIV<sup>2</sup> | 0.0320 | 0.0119 | 2.697 | < 0.01 |
| *Oga*<sup>K885N/K885N</sup> × DIV<sup>2</sup> | 0.0215 | 0.0117 | 1.846 | ns |
| *Oga*<sup>K885N/+</sup> × DIV<sup>3</sup> | -0.0008 | 0.0003 | -2.512 | < 0.05 |
| *Oga*<sup>K885N/K885N</sup> × DIV<sup>3</sup> | -0.0004 | 0.0032 | -1.323 | ns |
| Firing rate ~ Genotype × DIV(polynomial, 2) + Amplitude (scaled 0,1) |  |  |  |  |
|  | Estimate (β) | S.E. | z-value | p |
| Intercept | -0.6510 | 0.1495 | -4.35 | < 0.001 |
| *Oga*<sup>K885N/+</sup> | -0.1300 | 0.1837 | -0.71 | ns |
| *Oga*<sup>K885N/K885N</sup> | 0.0999 | 0.2243 | 0.45 | ns |
| DIV<sup>1</sup> | -0.0653 | 0.0223 | -2.93 | < 0.01 |
| DIV<sup>2</sup> | 0.0034 | 0.0009 | 3.81 | < 0.001 |
| Amplitude (scaled) | 0.2369 | 0.0061 | 39.09 | < 0.001 |
| *Oga*<sup>K885N/+</sup> × DIV<sup>1</sup> | 0.0136 | 0.0290 | 0.47 | ns |
| *Oga*<sup>K885N/K885N</sup> × DIV<sup>1</sup> | -0.0338 | 0.0328 | -1.03 | ns |
| *Oga*<sup>K885N</sup> × DIV<sup>2</sup> | 0.0008 | 0.0012 | 0.66 | ns |
| *Oga*<sup>K885N/K885N</sup> × DIV<sup>2</sup> | 0.0028 | 0.0013 | 2.16 | < 0.05 |
| Log(ISI) ~ Genotype × DIV + Amplitude |  |  |  |  |
|  | Estimate (β) | S.E. | t-value | p |
| Intercept | 6.0850 | 0.2450 | 24.832 | < 0.001 |
| *Oga*<sup>K885N/+</sup> | -1.3950 | 0.2137 | -6.525 | < 0.001 |
| *Oga*<sup>K885N/K885N</sup> | -1.6280 | 0.2697 | -6.038 | < 0.001 |
| DIV<sup>1</sup> | -0.2752 | 0.0250 | -11.024 | < 0.001 |
| DIV<sup>2</sup> | 0.0072 | 0.0010 | 7.45 | < 0.001 |
| Amplitude | -0.0012 | 0.0001 | -13.036 | < 0.001 |
| *Oga*<sup>K885N/+</sup> × DIV<sup>1</sup> | -0.1816 | 0.0324 | 5.608 | < 0.001 |
| *Oga*<sup>K885N/K885N</sup> × DIV<sup>1</sup> | -0.2100 | 0.0389 | 5.405 | < 0.001 |
| *Oga*<sup>K885N/+</sup> × DIV<sup>2</sup> | -0.0049 | 0.0013 | -3.843 | < 0.001 |
| *Oga*<sup>K885N/K885N</sup> × DIV<sup>2</sup> | -0.0054 | 0.0015 | -3.661 | < 0.001 |

**Supplementary Table 3:** List of proteins with associated tags and construct boundaries.

| **Plasmid** | **Protein** | **Tag** | **Antibody resistance** | **Boundaries** | **Mutations** |
| --- | --- | --- | --- | --- | --- |
| Hm hi13 3 | WT OGA N-term | His | Ampicillin | 11-396aa | - |
| L2P LEND | WT OGA C-term HAT | - | Kanamycin | 535aa-End | - |
| Hm A171 | OGA catalytic deficient mutant | His | Ampicillin | 11-396aa | D175N |
| L2P 85HE4 | OGA K885N mutant | - | Kanamycin | 535aa-End | K885N |

**Supplementary Table 4:**
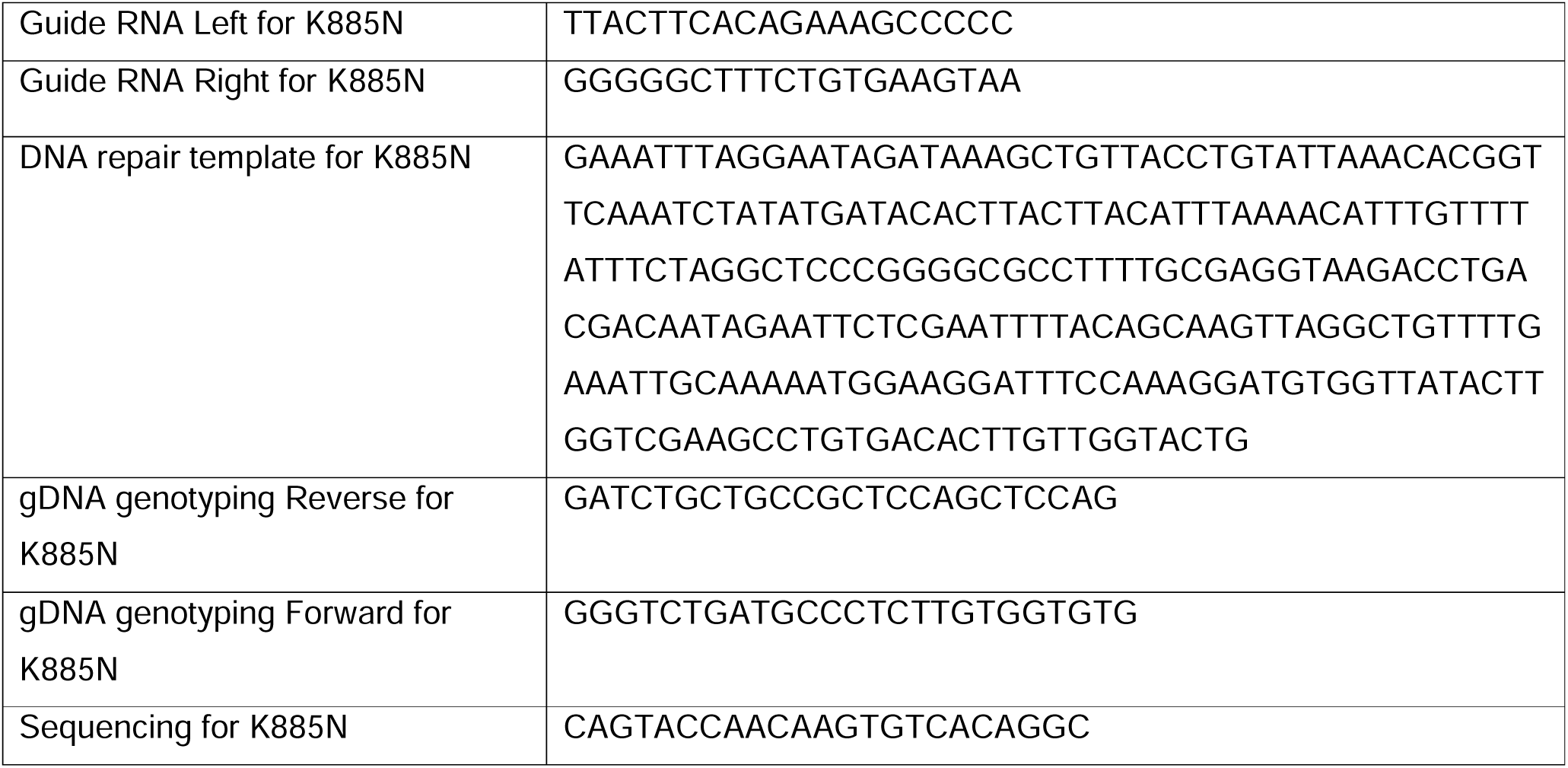
Sequences of reagents used for introducing the K885N mutation to *Oga* gene and genotyping of the mESCs and mice.

**Supplementary Table 5:**
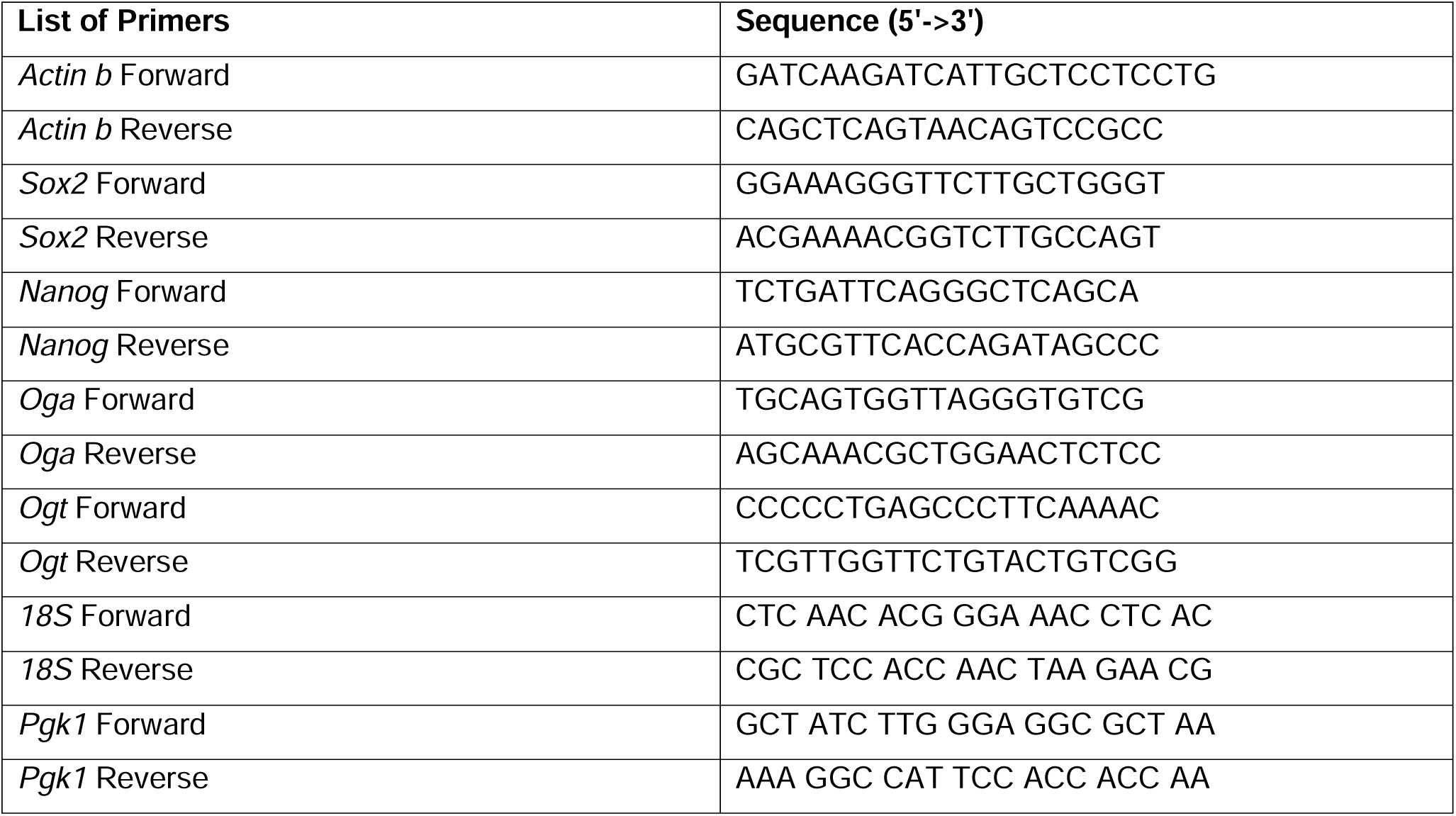
Sequences of primers used for qPCR analysis.

**Supplementary Table 6:** Numbers of wells and active channels recorded with HD-MEA. One well = one animal. Data was randomly down sampled to a maximum of 2000 active channels per Genotype-DIV combination.

| Genotype | DIV | Wells | N Active channels | N Active channels (down sampled) | N Spikes | N Spikes (downsampled) |
| --- | --- | --- | --- | --- | --- | --- |
| *Oga*<sup>+/+</sup> | 7 | 5 | 735 | 735 | 8593 | 8593 |
|  | 11 | 5 | 6797 | 2000 | 75991 | 31634 |
|  | 14 | 11 | 20703 | 2000 | 299585 | 75236 |
|  | 17 | 10 | 24160 | 2000 | 418318 | 80354 |
| *Oga*<sup>K855N/+</sup> | 7 | 16 | 2718 | 2000 | 29321 | 13564 |
|  | 11 | 16 | 16407 | 2000 | 258218 | 68613 |
|  | 14 | 16 | 34545 | 2000 | 642484 | 115571 |
|  | 17 | 12 | 42526 | 2000 | 1114013 | 181594 |
| *Oga*<sup>K885N/K885N</sup> | 7 | 3 | 441 | 441 | 3099 | 2644 |
|  | 11 | 3 | 2394 | 2000 | 39960 | 37291 |
|  | 14 | 3 | 3910 | 2000 | 76841 | 62107 |
|  | 17 | 2 | 5900 | 2000 | 159118 | 113457 |

